# How did the COVID-19 pandemic affect access to condoms, chlamydia and HIV testing, and cervical cancer screening at a population level in Britain? (Natsal-COVID)

**DOI:** 10.1101/2022.04.29.22274486

**Authors:** Emily Dema, Pam Sonnenberg, Jo Gibbs, Anne Conolly, Malachi Willis, Julie Riddell, Raquel Boso Perez, Andrew J. Copas, Clare Tanton, Chris Bonell, Clarissa Oeser, Soazig Clifton, Magnus Unemo, Catherine H Mercer, Kirstin R Mitchell, Nigel Field

## Abstract

**Objectives:** To investigate how differential access to key interventions to reduce sexually transmitted infections (STI), HIV, and their sequelae changed during the COVID-19 pandemic.

**Methods:** British participants (18-59y) completed a cross-sectional web survey one year (March to April 2021) after the initial lockdown in Britain. Quota-based sampling and weighting resulted in a quasi-representative population sample. We compared Natsal-COVID data with Natsal-3, a household-based probability sample cross-sectional survey (16-74y) conducted in 2010-12. Reported unmet need for condoms because of the pandemic and uptake of chlamydia testing/HIV testing/cervical cancer screening were analysed among sexually-experienced participants (18-44y) (n=2869, Natsal-COVID; n=8551, Natsal-3). Odds ratios adjusted for age (aOR) and other potential confounders (AOR) describe associations with demographic and behavioural factors.

**Results:** In 2021, 6.9% of women and 16.2% of men reported unmet need for condoms because of the pandemic. This was more likely among participants: aged 18-24 years, of Black or Black British ethnicity, and reporting same-sex sex (past five years) or one or more new relationships (past year). Chlamydia and HIV testing were more commonly reported by younger participants, those reporting condomless sex with new sexual partners, and men reporting same-sex partners; a very similar distribution to 10 years previously (Natsal-3). However, there were differences during the pandemic, including stronger associations with chlamydia testing for men reporting same-sex partners; with HIV testing for women reporting new sexual partners; and with cervical screening among smokers.

**Conclusions:** Our study suggests differential access to key primary and secondary STI/HIV prevention interventions continued during the first year of the COVID-19 pandemic. However, the available evidence does not suggest substantial changes in inequalities in since 2010–12. While the pandemic might not have exacerbated inequalities in access to primary and secondary prevention, it is clear that large inequalities persisted, typically among those at greatest STI/HIV risk.

**Key Messages:**

- Many MSM, people of Black ethnicity and young people (i.e. groups most impacted by STIs) reported unmet need for condoms because of the pandemic
- We compared inequalities in access to key interventions using Natsal-COVID (2021) and Natsal-3 (2010-12).
- During the pandemic (Natsal-COVID), there were stronger associations with chlamydia testing for MSM and with HIV testing for women reporting new sexual partners.
- There were stronger associations with cervical screening among smokers during the pandemic compared to 2010-12 (Natsal-3).
- However, we did not find strong evidence that vulnerable groups were at additional risk during the pandemic when compared to 2010-12.

## Introduction

Primary and secondary prevention methods aim to interrupt the transmission or consequences of sexually transmitted infections (STI) and HIV. For STIs and HIV, primary prevention aims to prevent infection occurring at all (e.g. condoms), while secondary prevention involves detection and treatment of infection before disease manifestations (e.g. testing for and treating early chlamydia/HIV infection or cervical cancer screening).^1^ Such interventions remained important during the COVID-19 pandemic because potentially risky sexual activity continued despite lockdowns,^2^ and STI/HIV diagnoses regained near pre-pandemic levels by the end of 2020.^3^ We know that different population groups experienced significant health inequalities during the pandemic due to the direct impacts of COVID-19, as well as impacts on the wider health system and society.^4^ We also know that there were significant pre-existing inequalities in uptake of SRH interventions and outcomes,^5-7^ and that the pandemic disrupted sexual and reproductive health (SRH) services, which is likely to have delayed diagnoses and led to worse outcomes. However, it is not known whether or how the pandemic affected these disparities in STI/HIV prevention.

In Britain, a national lockdown was announced on 23 March 2020, which lasted approximately four months and caused the most severe disruption. Restrictions on social and physical interactions continued throughout 2020. Another four-month national lockdown began in early January 2021. During this period, sexual health services reduced face-to-face consultations and prioritised vulnerable populations and symptomatic patients.^8^ Concerns about SARS-CoV-2 infection might also have decreased health-seeking behaviour.^9^

The National Survey of Sexual Attitudes and Lifestyles (Natsal)-COVID web-panel study was conducted to understand the population-level impact of the COVID-19 pandemic on SRH in Britain. Survey Wave 1 of Natsal-COVID was conducted four months (July-August 2020) after the announcement of the first national lockdown to understand initial changes in SRH service use.^10 11^ We observed that STI services were most likely to reach those most at-risk for STIs in those first four months, though there were often difficulties in access.^10 12^ Survey Wave 2, conducted a year after the initial lockdown captured key annual STI outcomes, such as HIV and chlamydia testing.^13^ Elsewhere, we have reported a reduction in chlamydia testing compared with Natsal-3, a household-based representative probability sample survey of the British population conducted from 2010 to 2012, while HIV testing and STI-related service use were similar to Natsal-3.^14^

In this paper, we investigated whether underlying inequalities in the uptake of key STI/HIV interventions might have increased during the first year of the pandemic. We used Natsal-COVID survey Wave 2 data on reported unmet need for condoms, chlamydia and HIV testing, and cervical cancer screening to assess the distribution in the general population and among groups experiencing a disproportionate burden of diagnoses (including men who have sex with men (MSM), young people, and people of Black ethnicity)^15^. We compared these distributions with data from Natsal-3 (2010-12) as the most recent representative population survey on sexual health in Britain. We hypothesized that differential access to key STI interventions has been exacerbated due to the pandemic.

## Methods

### Natsal-COVID Wave 2 study design

The Natsal-COVID survey Wave 2 was a quasi-representative web-panel survey of sexual health conducted one year after the first national COVID-19 lockdown in Britain. Data were collected using a short online questionnaire (median completion time: 13 minutes) through survey research company Ipsos-MORI’s web-panel. Participants were asked about uptake of STI interventions in the one year from 23 March 2020. The sample comprised longitudinal participants, who completed wave 1, and new cross-sectional participants recruited at Wave 2. The full questionnaire is available at https://www.natsal.ac.uk/natsal-covid-study. Details of the Natsal-COVID methods have been described elsewhere.^13^

### Participants and procedures of Natsal-COVID Wave 2

In total, 6,658 participants completed the survey, including 2,098 who also participated in wave 1. Data collection took place from 27 March to 26 April 2021. To achieve a quasi-representative sample of the British general population, we used quotas for age, gender, region (based on Office for National Statistics 2019 mid-year estimates) and social grade (based on Census 2011 data), and weighted the data to match the distributions found in the general population for the quotas and for ethnicity and sexual identity. We obtained ethical approval from University of Glasgow MVLS College Ethics Committee (reference 20019174) and London School of Hygiene and Tropical Medicine Research Ethics Committee (reference 22565). An anonymised dataset will be deposited with the UK Data Service to accompany the Natsal-COVID survey wave 1 data (serial number SN 8865) and datasets from previous decennial Natsal surveys, including Natsal-3 (serial number SN7799).

### Comparison with Natsal-3

We compared our findings with data from the Natsal-3 survey. Natsal-3 (2010-12) used a multistage, clustered, and stratified probability sample design.^16^ Interviewers visited all sampled addresses, identified residents in the eligible age range (16–74 years), and randomly selected one individual to participate in the survey. Participants then completed the survey in their own homes through a combination of face-to-face interviews and a self-completion interview. Interviews lasted about one hour on average. Details of the Natsal-3 methods have been described elsewhere.^16^

### Statistical measures and analysis

We used Stata (version 16.1) complex survey analysis functions to incorporate weighting and stratification. Outcomes of interest, including question wording, are shown in Table 1.

**Table 1.**
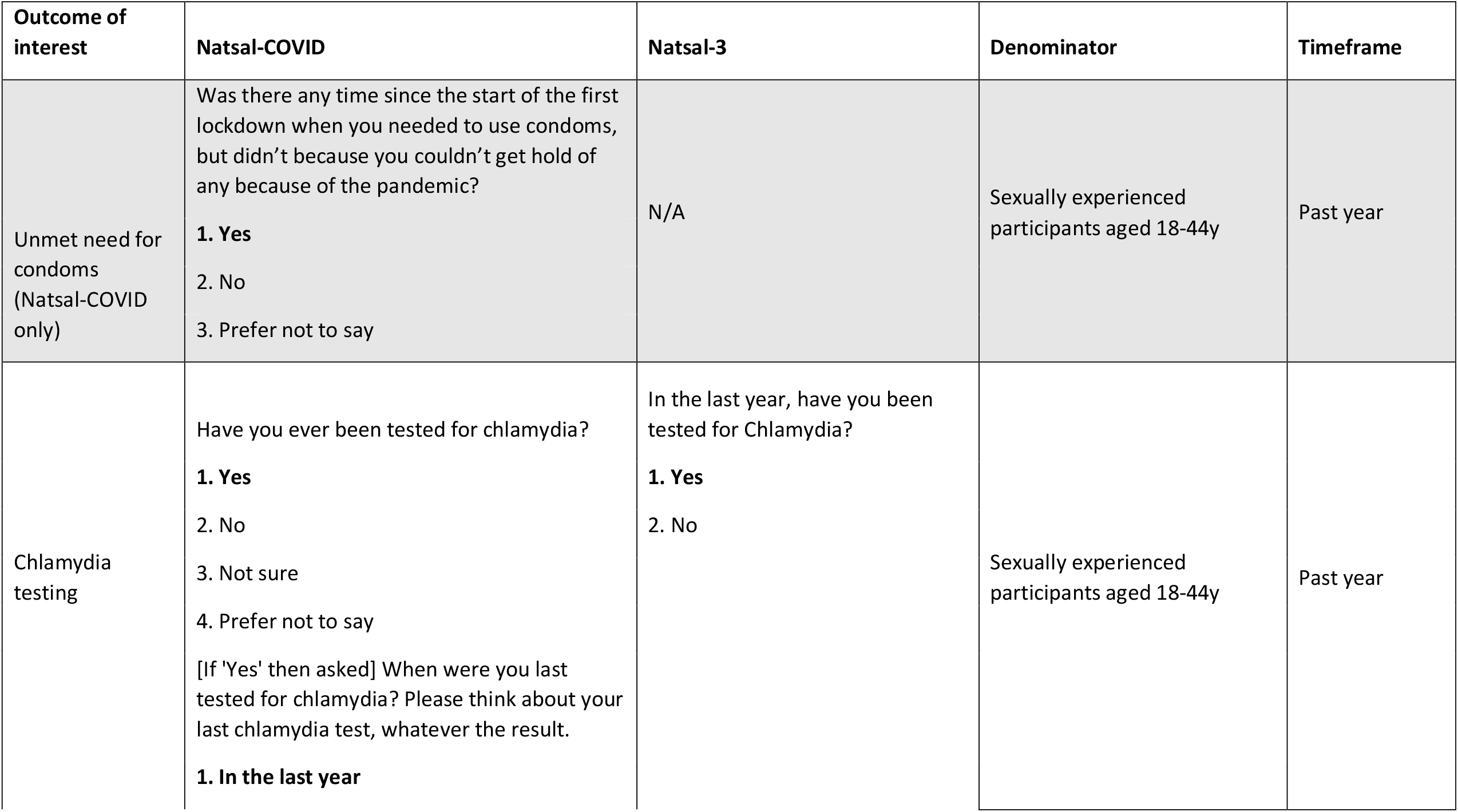
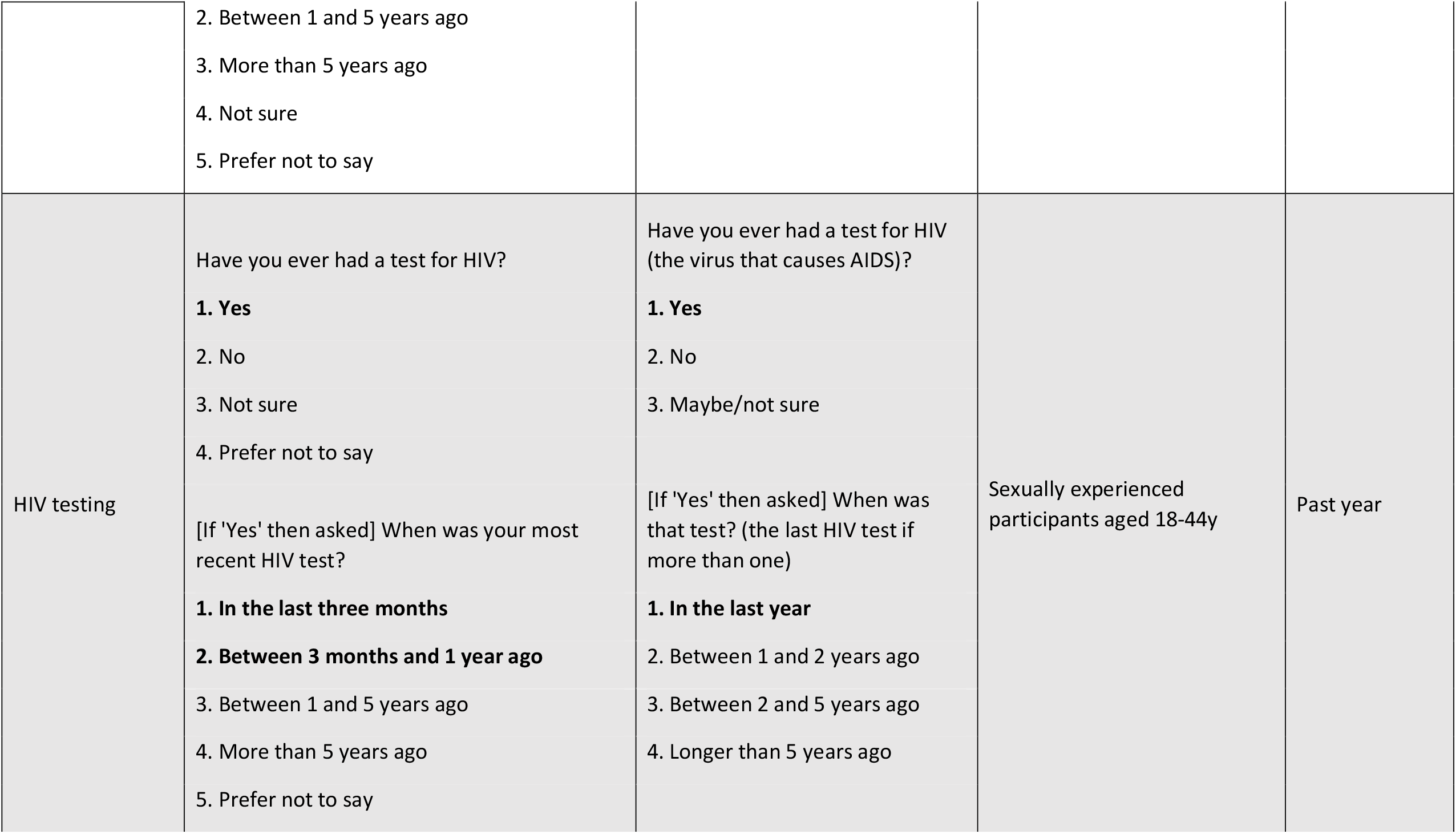
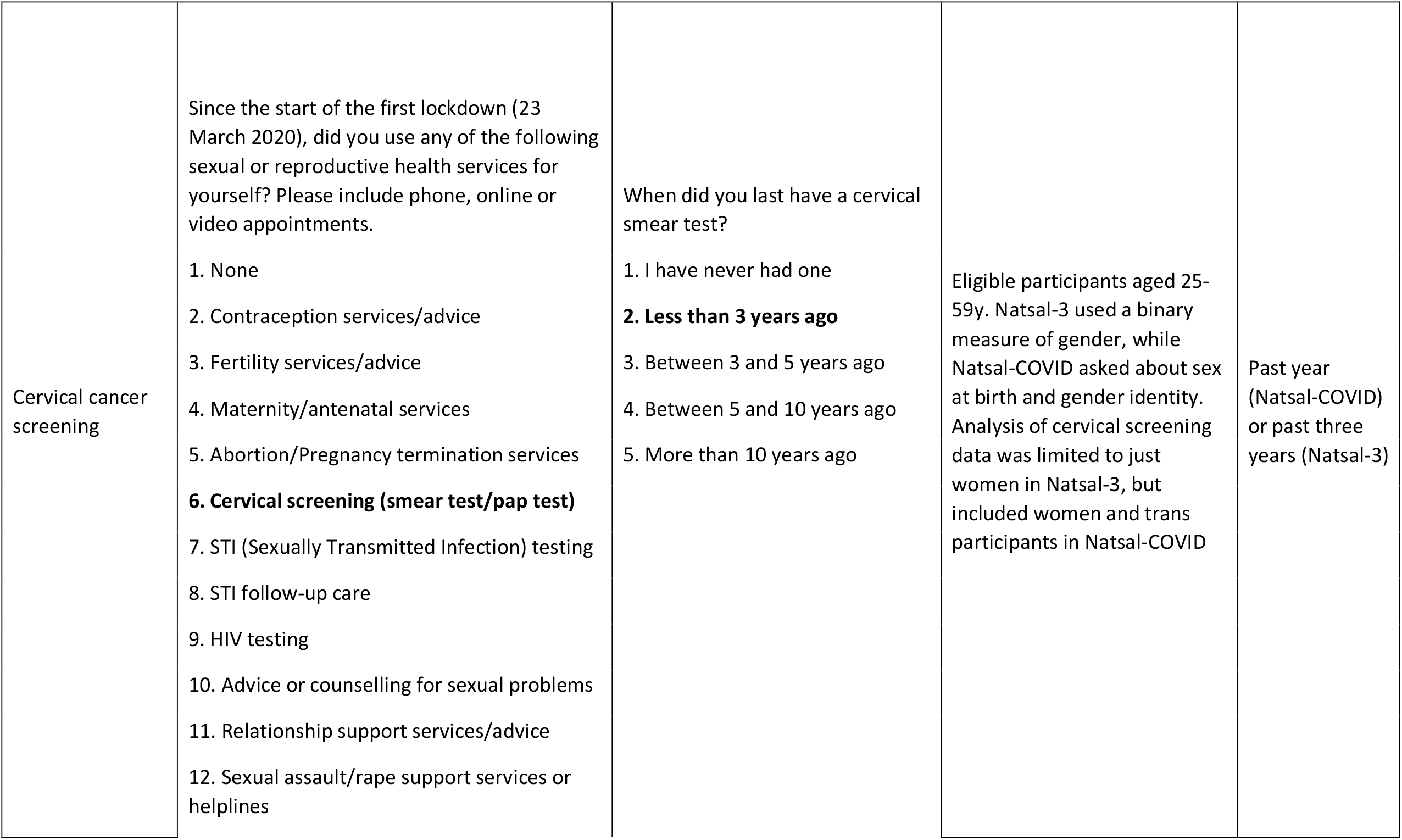
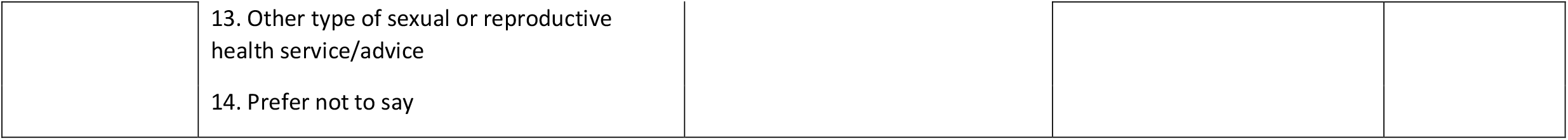
Outcomes of interest in Natsal-COVID and Natsal-3

| Outcome of interest | Natsal-COVID | Natsal-3 | Denominator | Timeframe |
| --- | --- | --- | --- | --- |
| Unmet need for condoms<br>(Natsal-COVID only) | Was there any time since the start of the first lockdown when you needed to use condoms, but didn't because you couldn't get hold of any because of the pandemic?<br>**1. Yes**<br>2. No<br>3. Prefer not to say | N/A | Sexually experienced participants aged 18-44y | Past year |
| Chlamydia testing | Have you ever been tested for chlamydia?<br>**1. Yes**<br>2. No<br>3. Not sure<br>4. Prefer not to say<br>[If 'Yes' then asked] When were you last tested for chlamydia? Please think about your last chlamydia test, whatever the result.<br>**1. In the last year** | In the last year, have you been tested for Chlamydia?<br>**1. Yes**<br>2. No | Sexually experienced participants aged 18-44y | Past year |
|  | 2. Between 1 and 5 years ago<br>3. More than 5 years ago<br>4. Not sure<br>5. Prefer not to say |  |  |  |
| HIV testing | Have you ever had a test for HIV?<br>**1. Yes**<br>2. No<br>3. Not sure<br>4. Prefer not to say<br>[If 'Yes' then asked] When was your most recent HIV test?<br>**1. In the last three months**<br>**2. Between 3 months and 1 year ago**<br>3. Between 1 and 5 years ago<br>4. More than 5 years ago<br>5. Prefer not to say | Have you ever had a test for HIV (the virus that causes AIDS)?<br>**1. Yes**<br>2. No<br>3. Maybe/not sure<br>[If 'Yes' then asked] When was that test? (the last HIV test if more than one)<br>**1. In the last year**<br>2. Between 1 and 2 years ago<br>3. Between 2 and 5 years ago<br>4. Longer than 5 years ago | Sexually experienced participants aged 18-44y | Past year |
| Cervical cancer screening | Since the start of the first lockdown (23 March 2020), did you use any of the following sexual or reproductive health services for yourself? Please include phone, online or video appointments.<br>1. None<br>2. Contraception services/advice<br>3. Fertility services/advice<br>4. Maternity/antenatal services<br>5. Abortion/Pregnancy termination services<br>**6. Cervical screening (smear test/pap test)**<br>7. STI (Sexually Transmitted Infection) testing<br>8. STI follow-up care<br>9. HIV testing<br>10. Advice or counselling for sexual problems<br>11. Relationship support services/advice<br>12. Sexual assault/rape support services or helplines | When did you last have a cervical smear test?<br>1. I have never had one<br>**2. Less than 3 years ago**<br>3. Between 3 and 5 years ago<br>4. Between 5 and 10 years ago<br>5. More than 10 years ago | Eligible participants aged 25-59y. Natsal-3 used a binary measure of gender, while Natsal-COVID asked about sex at birth and gender identity. Analysis of cervical screening data was limited to just women in Natsal-3, but included women and trans participants in Natsal-COVID | Past year (Natsal-COVID) or past three years (Natsal-3) |

|  |  |
| --- | --- |
|  | 13. Other type of sexual or reproductive health service/advice |
|  | 14. Prefer not to say |

Data from Natsal-COVID are presented for all participants and separately for men (including trans men) and women (including trans women). While we did not present estimates for participants who identified ‘in another way’ for gender, these 22 participants were included in estimates presented for ‘all’. For analysis of cervical screening, we included all participants described female at birth, which included some trans men and non-binary people. Natsal-3 used a binary measure of gender.

We examined the outcome of ‘unmet need for condoms’ among sexually-experienced participants (i.e., any lifetime vaginal, anal, oral sex or other genital contact) by asking “Was there any time since the start of the first lockdown when you needed to use condoms, but didn’t because you couldn’t get hold of any because of the pandemic?” Participants aged 45-59 years were excluded from this analysis due to the low burden of STIs in this age group. Of the 6658 Natsal-COVID participants aged 18–59 years, 4323 were aged 18–44 years, of whom 3869 were sexually-experienced and included in analysis. Although some sexually-experienced participants (n=270 men and n=240 women) did not report any sexual partners in the past year, they were still included in denominators for ‘unmet need for condoms’ since disrupted access to condoms might have prevented some participants from having sex. This question was not asked in Natsal-3.

We estimated reported chlamydia and HIV testing in the past year among sexually-experienced participants (18–44y) for Natsal-COVID and Natsal-3. Natsal-3 participants reporting at least one lifetime sexual partner were considered ‘sexually-experienced’. Of 15,162 Natsal-3 participants, 8969 were aged 18–44 years, and, of these, 8551 were sexually-experienced and included in this analysis.

We estimated self-reported cervical cancer screening among all eligible participants (i.e., reported being described female at birth (Natsal-COVID) or women (Natsal-3) and aged 25– 59 years). This age group was chosen to most closely reflect the UK national screening programme eligibility of age 25–64 years. Cervical screening estimates are presented for eligible participants aged 25-59 years for the past year (Natsal-COVID) or past three years (Natsal-3); therefore, we focused on comparing characteristics associated with the uptake of cervical screening between the two surveys, rather than comparing prevalence estimates.

Men who have sex with men (MSM) in Natsal-COVID and Natsal-3 were defined as men (based on self-reported gender identity in Natsal-COVID) reporting at least one same-sex partner (as defined by participant) in the past five years.

We used logistic regression to calculate age-adjusted odds ratios (aORs) for all outcomes to investigate how access to STI/HIV interventions varied by sociodemographic and behavioural factors. For ‘unmet need for condoms’, we also produced odds ratios adjusted by age, sociodemographic, and behavioural variables. To establish independent associations with ‘unmet need for condoms’, the model was adjusted for sociodemographic (age, region, rurality, ethnicity, and relationship formation) and behavioural (sexual partners in the past year and previous same-sex experience in the past five years) factors. Where possible, we compared aORs in Natsal-COVID analyses with those generated from Natsal-3 data to investigate whether patterns of association differed between these studies. We describe the differences in the strength of associations and test for differences in the distribution of associations by including interaction terms in the regression models.

### Patient and Public Involvement statement

Patients or the public were not directly involved in the design, conduct, reporting, or dissemination plans of the Natsal-COVID study due to the urgency of the research during the pandemic. However, members of the public were involved in the design of the Natsal-4 questionnaire, upon which the Natsal-COVID questionnaire was based.

## Results

### Primary STI prevention

#### Unmet need for condoms

Among sexually-experienced participants (18–44y), 6.9% of women and 16.2% of men reported unmet need for condoms in the past year because of the pandemic (Table 2). Participants aged 18-24 years (women 16.8% and men 33.1%) and MSM (36.8%) were more likely to report this. Unmet need for condoms was even higher in young MSM (50.4% of 89 MSM aged 18–29 years old).

**Table 2.**
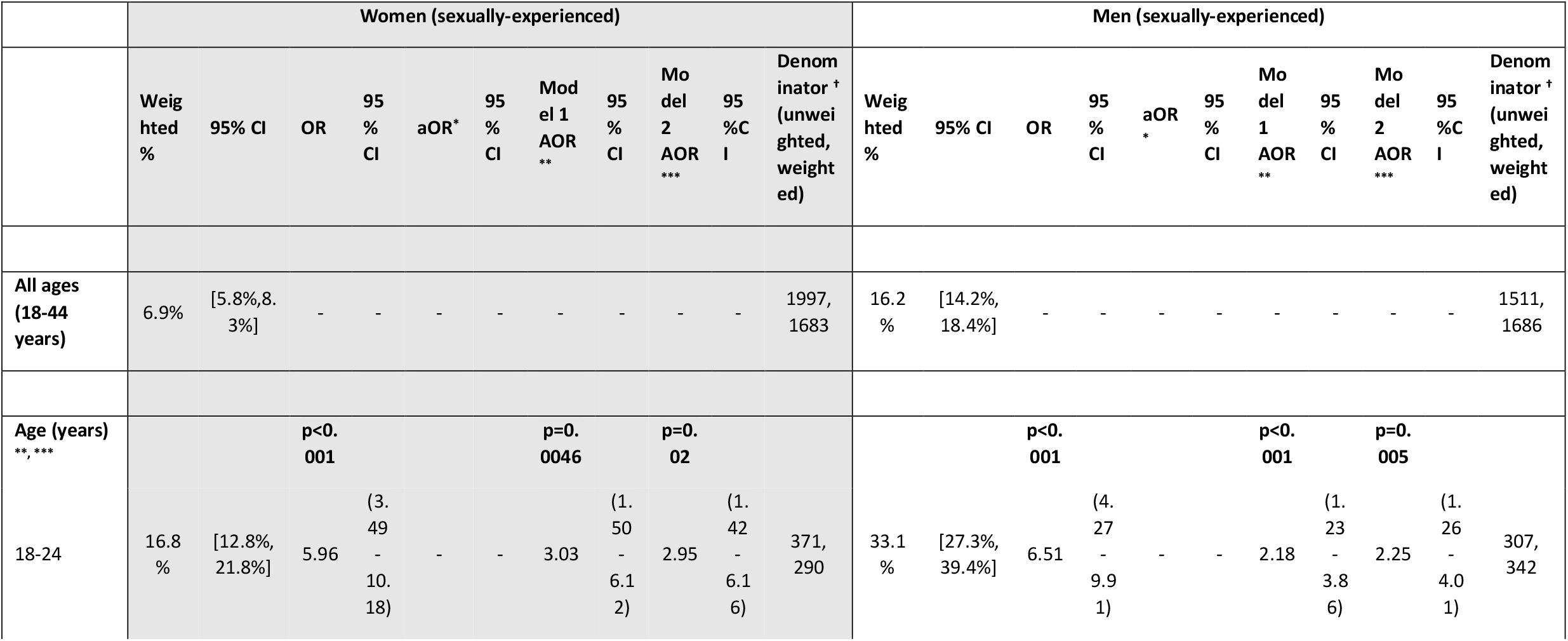
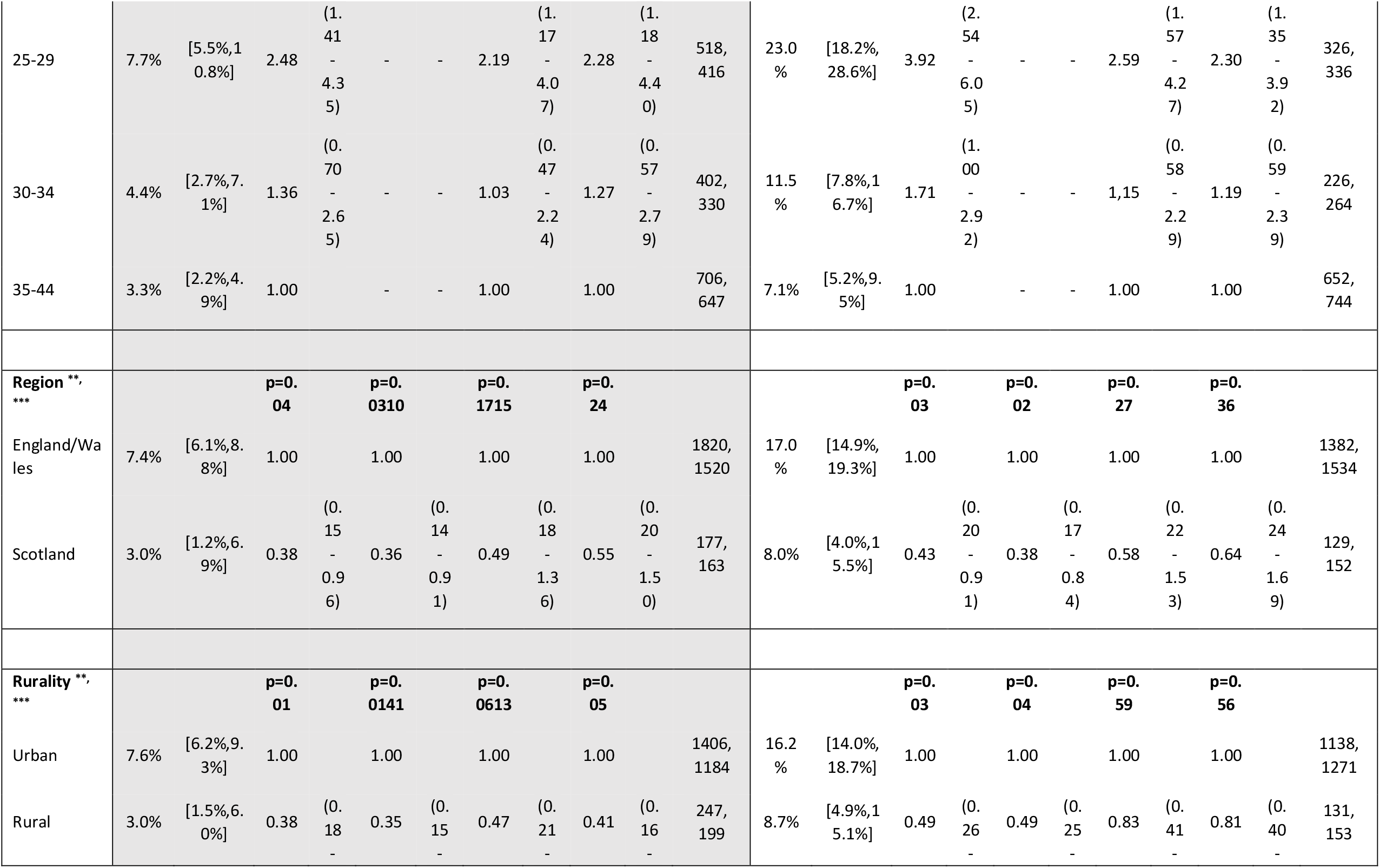
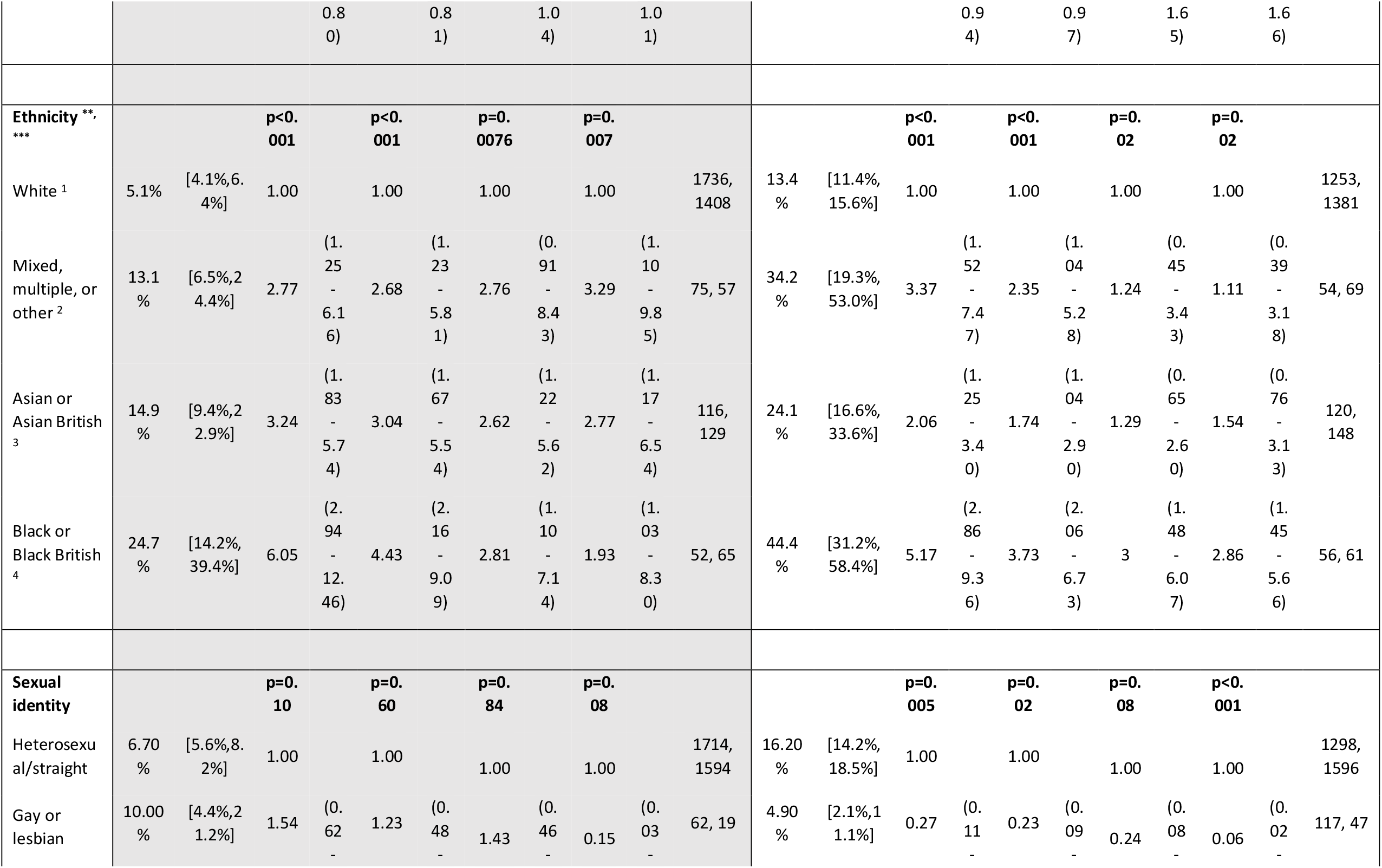
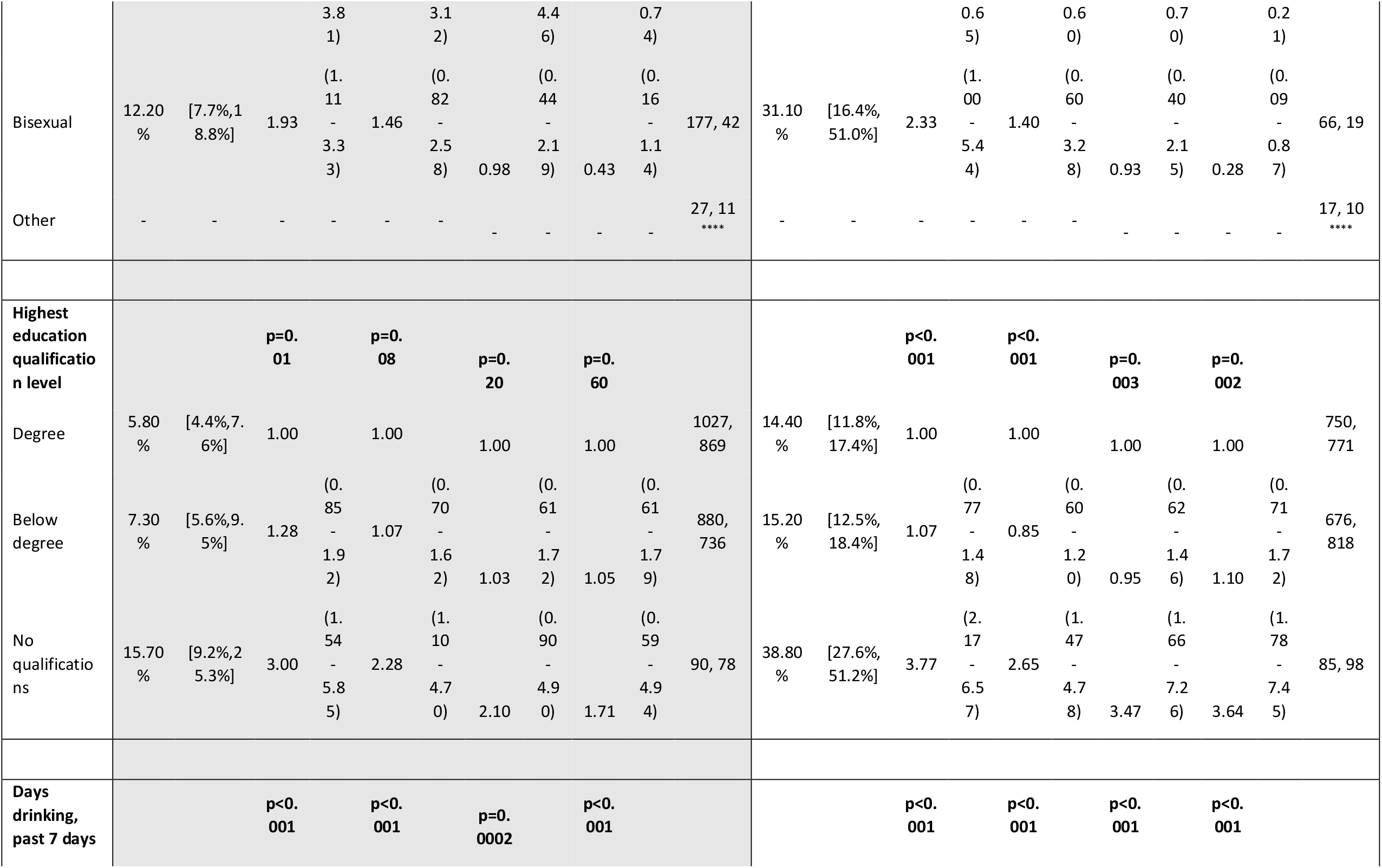
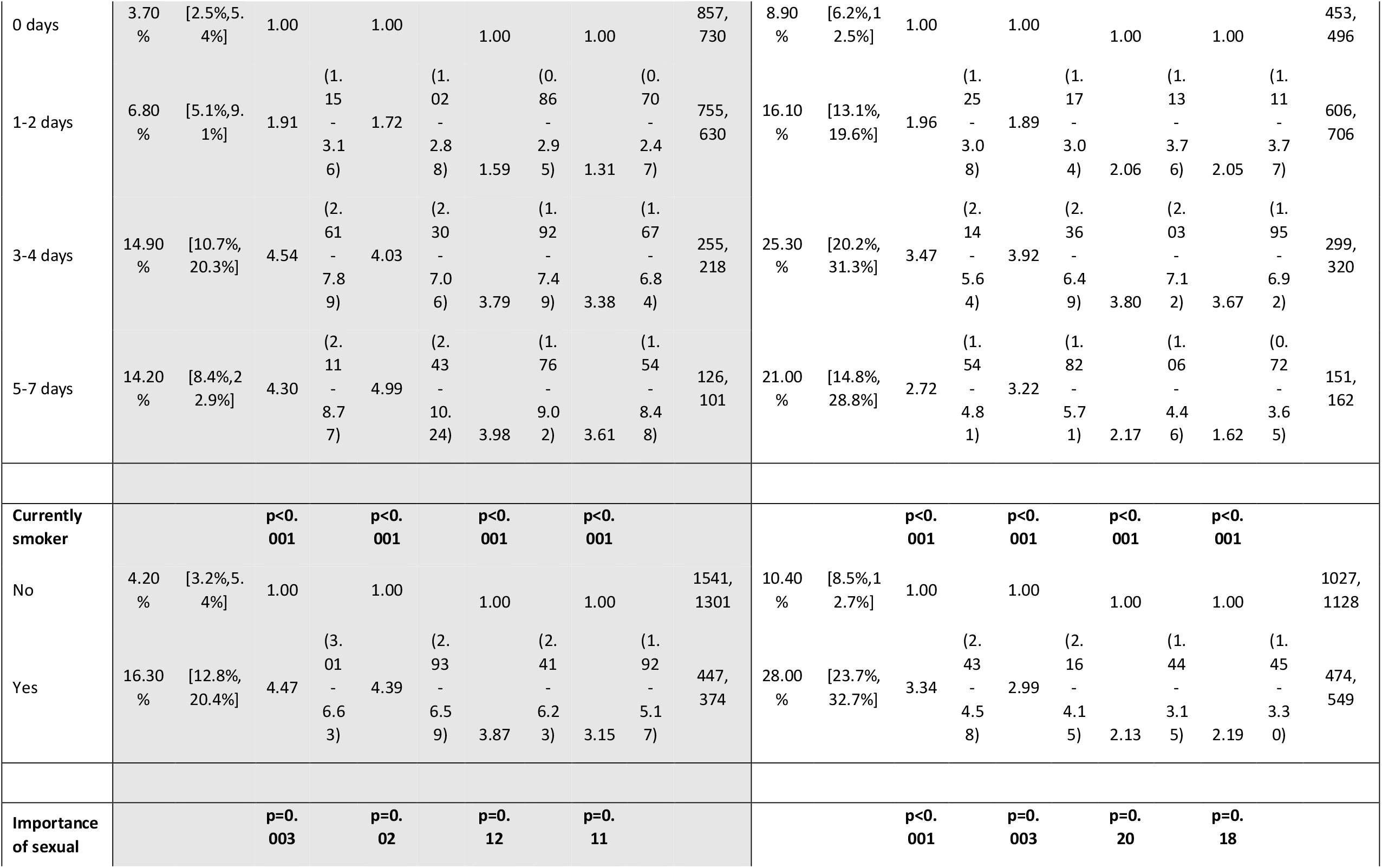
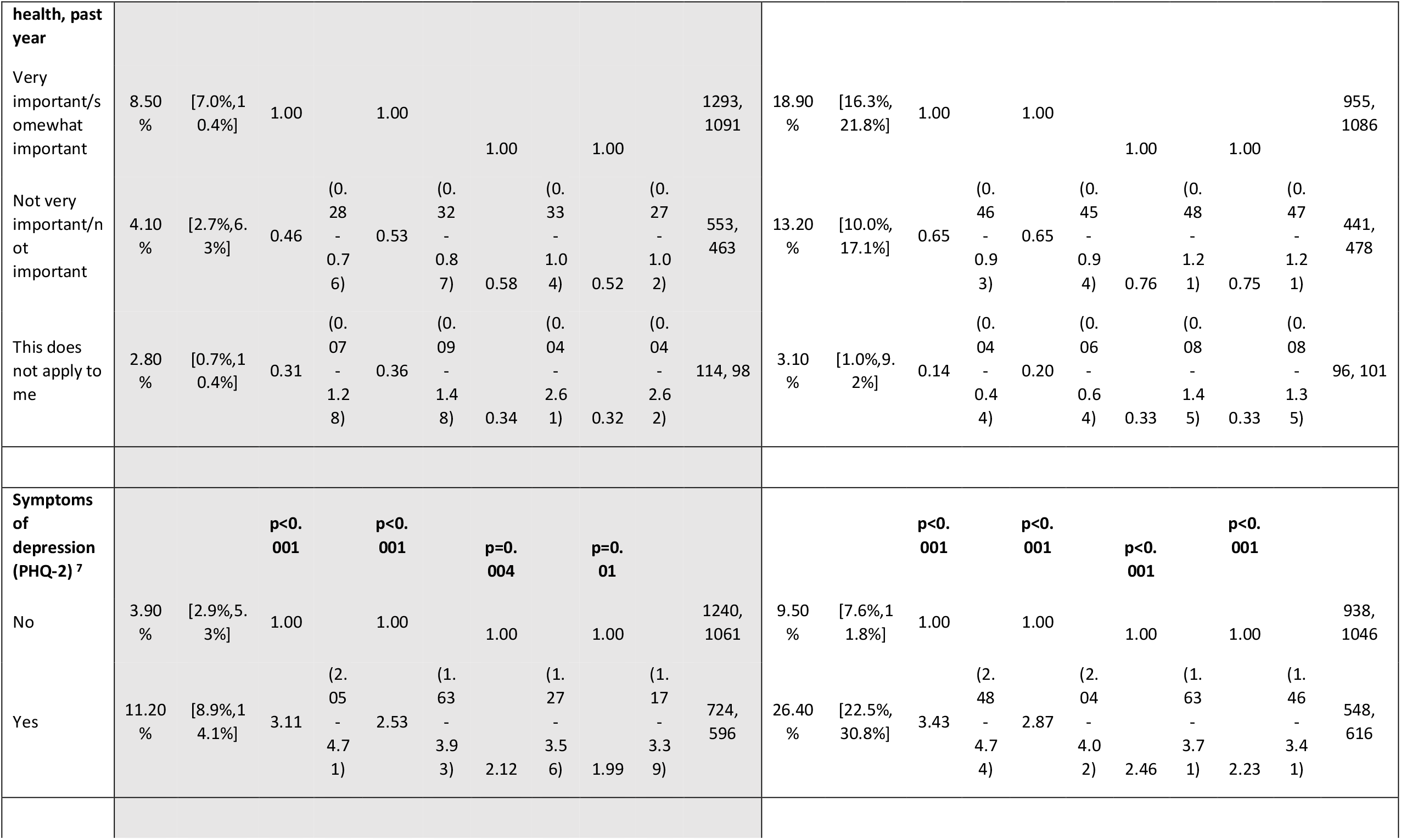
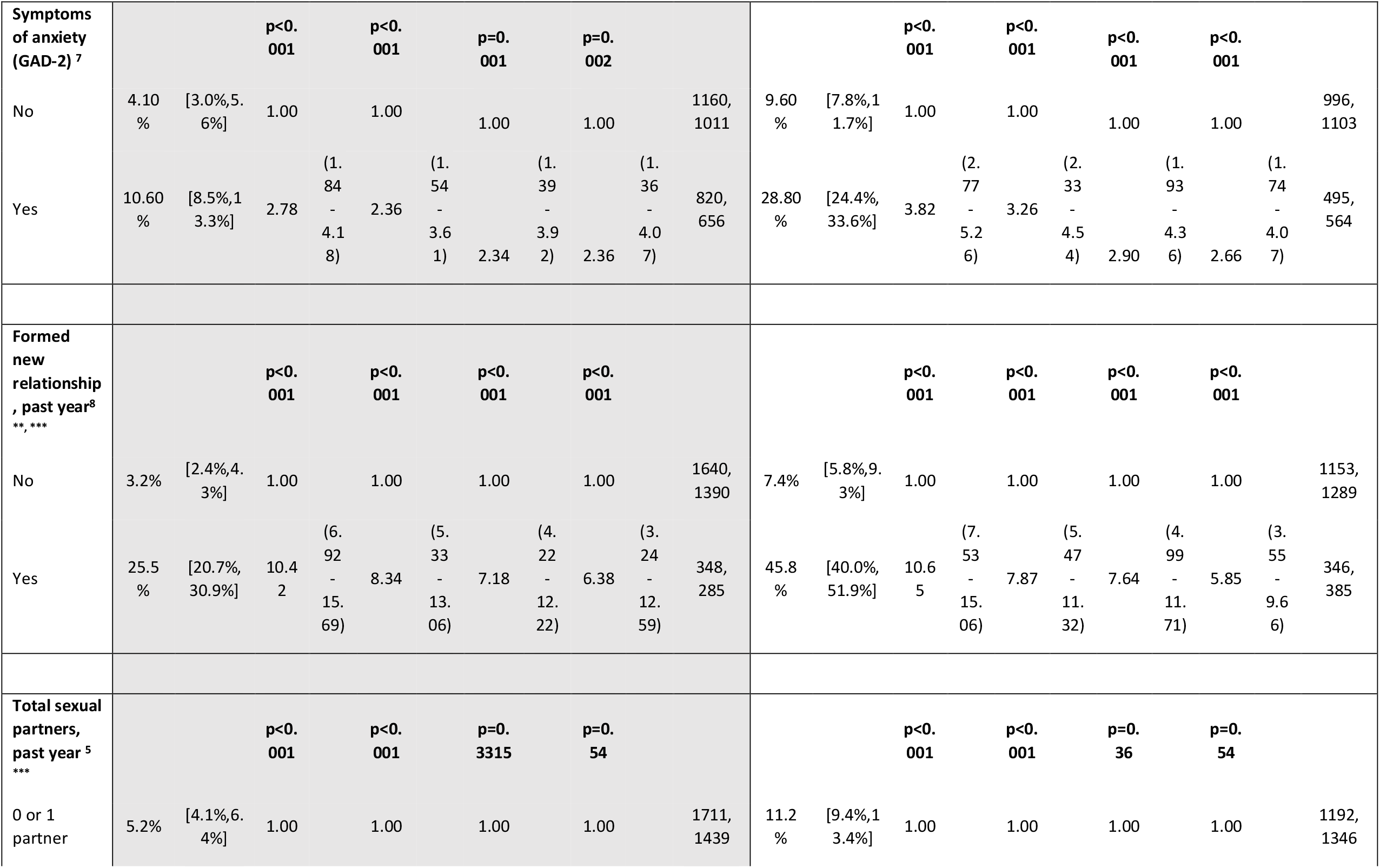
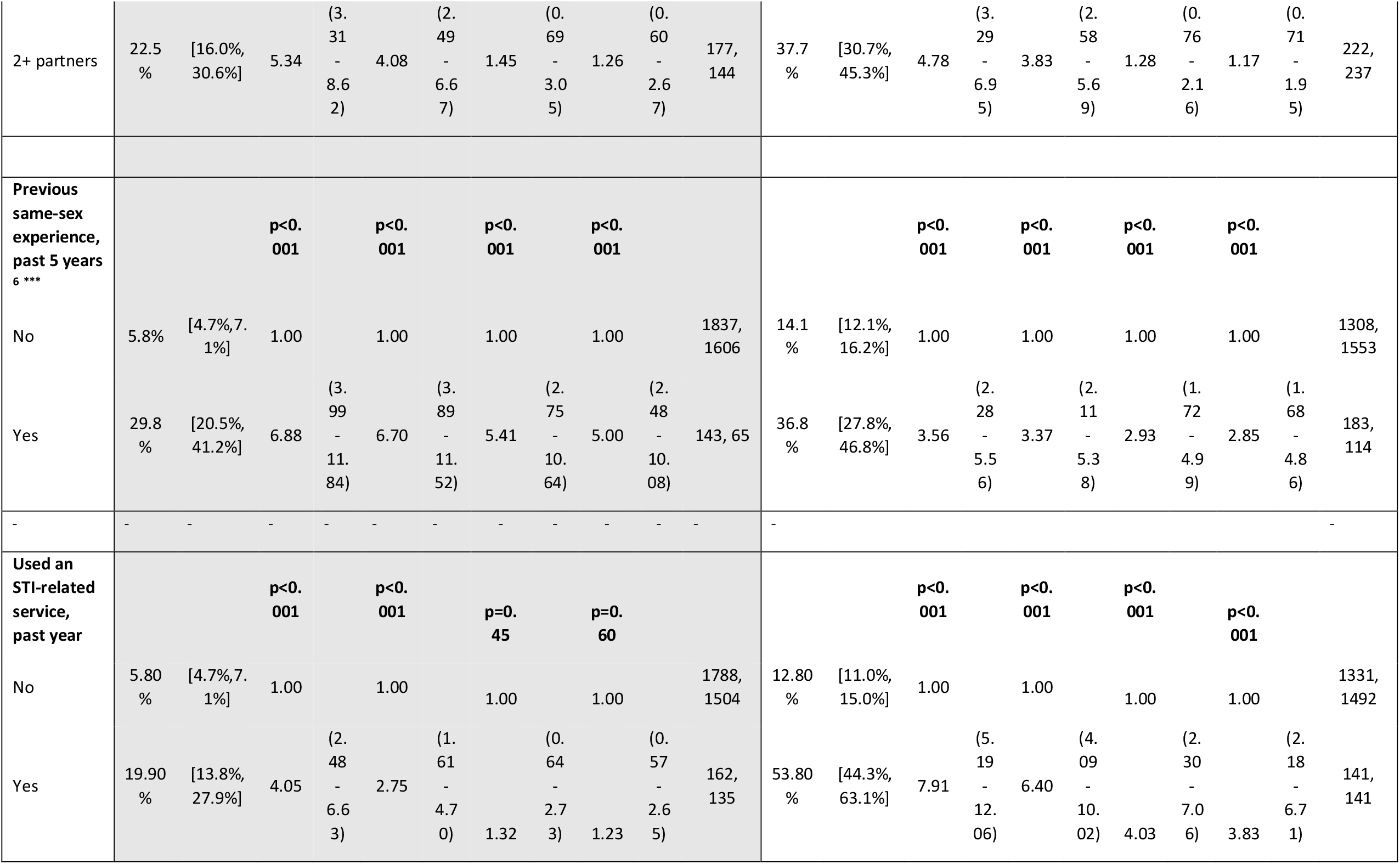
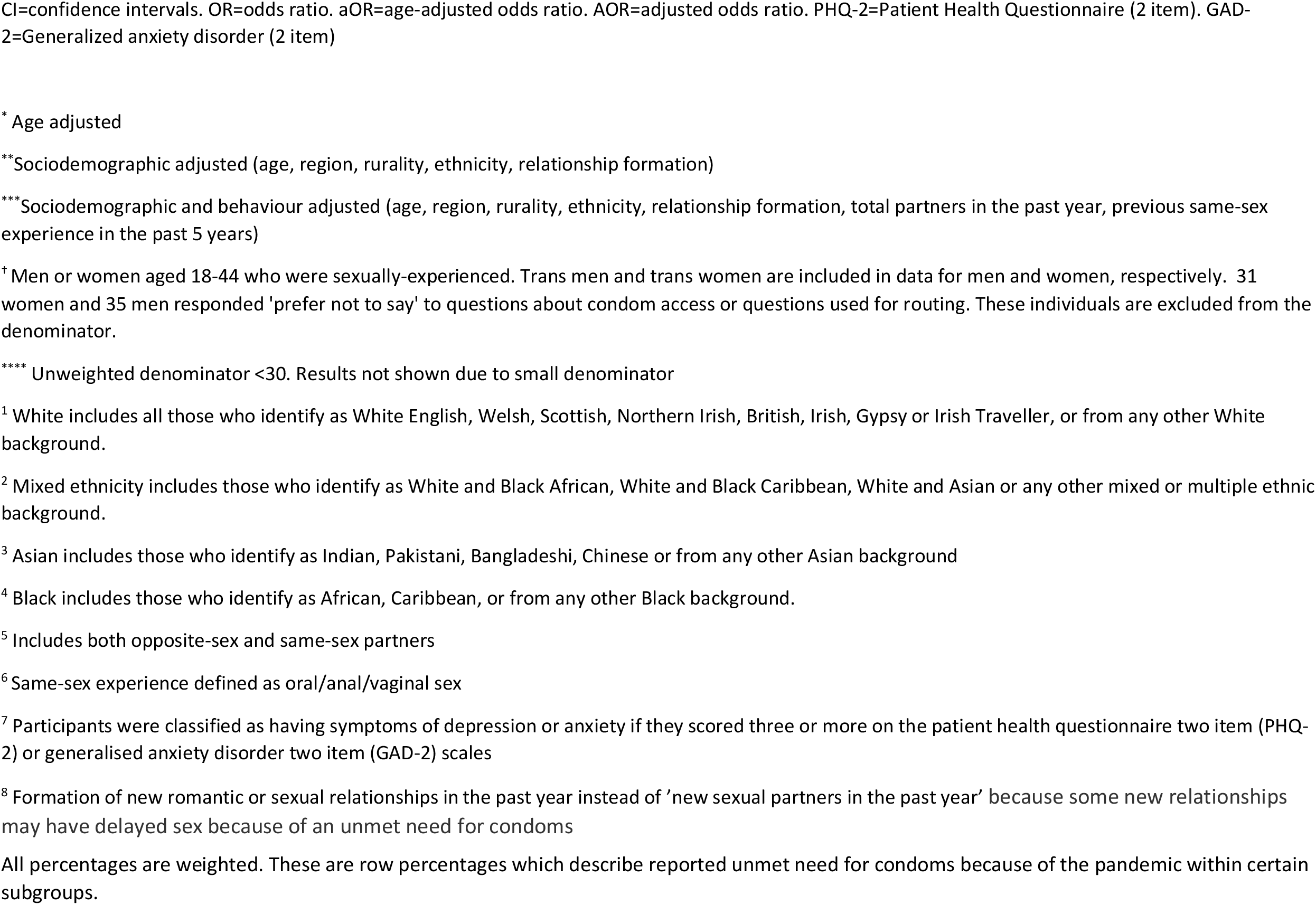
Variations in reporting unmet need for condoms because of the pandemic among sexually-experienced women and men aged 18-44 years in the first year following the start of a national lockdown in Britain (23/03/2020)

Using a regression model adjusted for sociodemographic and behavioural characteristics, unmet need for condoms because of the pandemic was most likely to be reported by younger participants and, among men, those identifying as Black or Black British (Table 2). Participants who reported symptoms of depression or anxiety were more likely to report unmet need for condoms.

There were also strong associations between unmet condom need and behavioural markers of HIV/STI risk. Participants who reported forming new relationships in the past year or a same-sex experience in the past five years were more likely to report unmet need for condoms (44.1% of women who reported previous same-sex experience also reported at least one opposite sex partner in the past five years). Among participants who reported unmet need for condoms, 47.0% (39.6–54.5%) of men and 34.4% (25.9–44.0%) of women also reported condomless sex on the first occasion with a new partner during the past year. By comparison, in the group that did not report unmet need for condoms, only 13.9% (11.7–16.4%) of men and 8.6% (7.3–10.2) of women reported condomless sex on the first occasion with a new partner (aOR for condomless with new partner: women, 4.42 (2.81–6.95); men, 4.67 (3.21– 6.78); data not shown). Among men but not women, participants who reported use of STI-related services in the past year were more likely to report unmet need for condoms in the adjusted model.

### Secondary STI prevention

#### Chlamydia and HIV testing

Among sexually-experienced participants (18-44y), 7.3% of women and 4.1% of men reported a chlamydia test in the past year, which was significantly lower than the proportions reported in Natsal-3 (2010–12) (25.1% women; 15.1% men). HIV testing in the past year was reported by 8.6% of women and 6.5% of men in Natsal-COVID Wave 2, similar to the 10.4% of women and 6.0% of men in Natsal-3. (Supplementary tables 1/2)

The direction and strength of associations for most independent variables with chlamydia and HIV testing were similar for Natsal-COVID and Natsal-3, based on interaction terms (Figure 1, Supplementary tables 1/2). For example, in both surveys, participants aged 18–24 were more likely to report an HIV test compared with those aged 35–44 years, Black or Black British participants were more likely to report testing than White participants, and MSM were more likely than other men to report testing. In each case, the odds ratios or strength of associations were very similar.

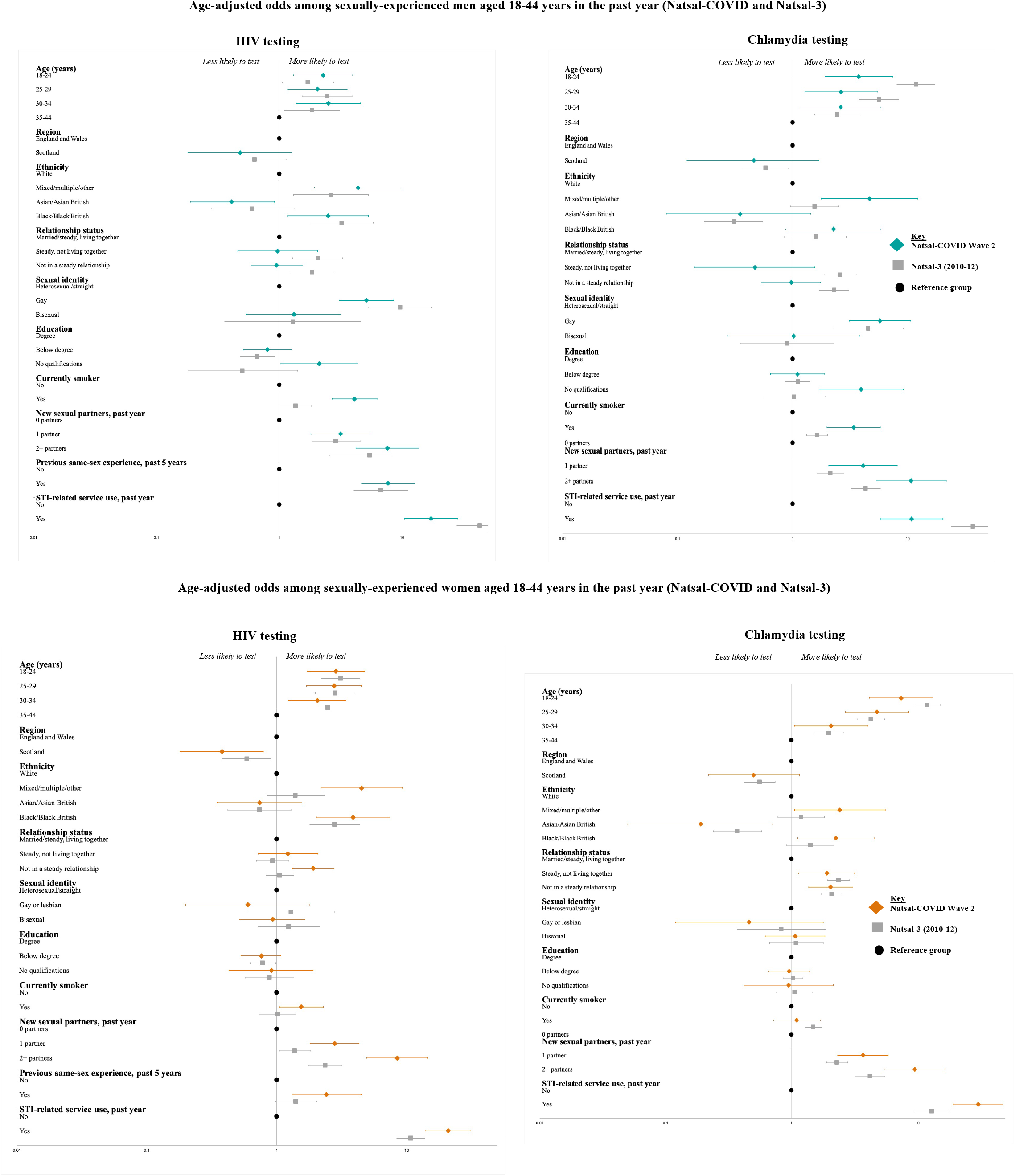

Nevertheless, there were some statistically significant interactions suggesting several differences between testing during the pandemic and 10 years previously. For example, young people (18–24y) were significantly more likely to report chlamydia testing compared with the oldest age group in both surveys (i.e., the direction of association was the same), and while the strength of this age association was similar for women across the surveys, it was significantly stronger for men in Natsal-3 than Natsal-COVID (interaction p=0.01). MSM were also more likely to report chlamydia testing in Natsal-COVID than Natsal-3 (interaction p=0.04).

#### Cervical cancer screening

Among participants described female at birth (25–59y) in Natsal-COVID, 10.3% reported use of cervical cancer screening services in the past year. In Natsal-3, 70.6% of women reported cervical screening in the past three years.

Again, associations for reported cervical screening were broadly similar to those in Natsal-3 (Figure 2, Supplementary table 3). The youngest participants (25–29y) were more likely to report screening compared with participants aged 44–59 years in both surveys, although the association with age was stronger in Natsal-COVID than Natsal-3 (interaction p=0.01). Gay or lesbian participants were less likely to screen than heterosexual participants in Natsal-COVID, while there was no association with sexual identity in Natsal-3 (interaction p=0.01). Notably, participants who reported smoking were more likely to report screening in Natsal-COVID, while this same group was less likely to screen in Natsal-3. Cervical screening was associated with markers of sexual risk. Participants in Natsal-COVID reporting two or more sexual partners in the past year were more likely to report screening compared to those with zero partners, but there was no association for this variable in Natsal-3 (interaction p=0.01).

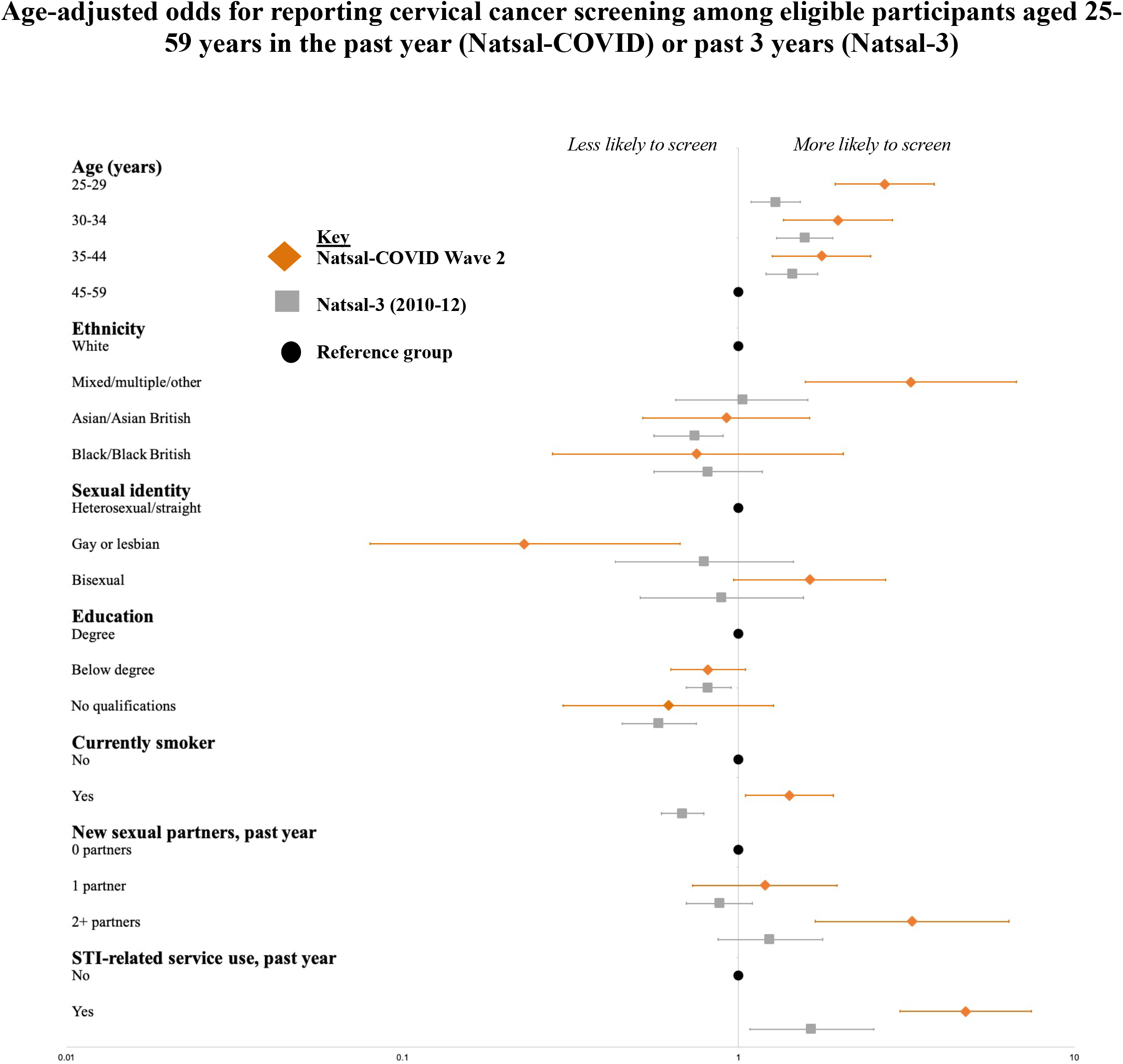

## Discussion

### Principal findings

Findings from this large, quasi-representative survey of the British population indicate there was differential access to key primary and secondary STI and HIV prevention interventions during the COVID-19 pandemic in Britain, particularly for young people, MSM, and those reporting new sexual partners. However, we did not find strong evidence that vulnerable groups were at additional risk during the pandemic when compared to 2010-12.

Regarding primary prevention, use of condoms is a highly cost-effective way to prevent transmission of STIs/HIV and unplanned pregnancy.^17^ However, 6.9% of women and 16.2% of men aged 18-44 years reported unmet need for condoms in the past year because of the pandemic. This was even higher for young men aged 18–24 years (33%) and MSM aged 18– 29 years (50%). Participants who reported one or no partners in the past year (i.e., at low risk for STIs) still reported an unmet need for condoms, which could indicate that some people were avoiding sex because they were unable to access condoms. It is also striking that participants reporting symptoms of depression or anxiety were more likely to report unmet need for condoms, though we are unable to determine causality. On the other hand, participants who reported unmet need for condoms were more likely to report sexual behaviours associated with STI/HIV risk. For example, they were more likely to report condomless sex with new partners, which suggests that improving access to condoms might support higher levels of condom use with new partners, in turn reducing STI/HIV transmission. Notably, many participants reporting unmet need for condoms also reported use of STI-related services in the past year, particularly among men, suggesting a role for SRH services in improving access to free or low-cost and easily accessible condoms, which may have been affected by remote service provision or other service changes. MSM, people of Black ethnicity and young people are amongst the groups most impacted by STIs in Britain,^15^ and it is concerning that a high proportion of individuals in these groups were unable to access condoms when they needed them. A Scottish web survey conducted in July 2020 corroborates our findings on unmet need for condoms, especially among young people ^18^. With advances in testing and treatment technologies, condoms are often forgotten in discourse about STI/HIV prevention. Anecdotal evidence suggests that provision of condoms at SRH services has reduced in the past decade and that remote service provision has further limited access to condoms during the pandemic. Given the high proportion of participants at most risk for STI and HIV transmission who reported unmet need for condoms in the past year because of the pandemic, improving accessibility to free or low-cost condoms in Britain should be prioritised.

The distribution in the population of reporting chlamydia and HIV testing were broadly similar for Natsal-COVID (2021) and Natsal-3 (2010-12). Key populations known to be at most risk for STI transmission, including young people, MSM and those reporting condomless sex with new partners, continue to be most likely to engage with sexual health services and the strengths of association between the different groups were similar in both surveys. In the past decade, HIV testing among MSM has increased due to targeted campaigns.^19^ However, we did not detect a stronger association with HIV testing among MSM in Natsal-COVID compared with Natsal-3—potentially due to a reversal of the upward trend in HIV testing among MSM in the years immediately prior to the pandemic.^19 20^

Although we were unable to directly compare population estimates because of differences in the reporting timeframes, patterns of reported access to cervical cancer screening were similar in Natsal-COVID and Natsal-3. However, there was higher reported use of screening services among younger participants (25–29 years) in Natsal-COVID, which might suggest either a longer-term trend over the past decade and/or a greater willingness to access services during the pandemic in younger compared to older participants, who might have perceived higher risk of severe COVID-19 disease. In Natsal-3, reported uptake of cervical cancer screening was lower among smokers, while this group was more likely to screen in Natsal-COVID. At a population level, smoking has declined substantially in the past decade, particularly among 18-24 year olds^21^, so women who smoke in 2021 might be a different group to those who smoked in 2010–12. Nevertheless, the finding that smokers were more likely to report cervical screening could be positive, given the additional risk for cervical cancer brought by smoking ^22^. Surveillance data suggest a decrease in invitations and screening in 2020 compared with the previous year, which corroborates Natsal-COVID Wave 1 and Wave 2 findings suggesting a potential backlog of need among those eligible for cervical screening.^10 14 23^

### Comparison with other studies

Reprioritisation of health care services, including SRH, due to COVID-19 led to unmet need,^10^ even though there was a reduction in new sexual partners, particularly among young people and MSM.^14^ Data from the UK Health Security Agency (UKHSA), formerly Public Health England, demonstrated a fall in bacterial STI testing from 2019 to 2020 among younger people (aged 15–25 years), people of Asian or Black ethnicity, and heterosexual men, though there was a small increase in testing among MSM.^24^ Surveillance data also showed that the burden of STIs remained greatest in those aged 15–24 years, as well as Black ethnic minorities and MSM in 2020.^15^

### Strengths and limitations

No previous study has examined whether differential access to key interventions to prevent STI or HIV and their sequelae might have changed at a population level due to the COVID-19 pandemic.^20^ However, our study also has limitations.^13^ Whilst it benefited from a questionnaire developed by the Natsal team to obtain high-quality data while navigating pandemic-related circumstances and used a large national sample, with quota sampling and weighting to improve generalisability, the Natsal-COVID study is not a probability sample. Specific prevalence estimates should be treated with particular caution given expected selection and response biases.

Due to the lack of population-level data on access to key STI/HIV prevention interventions by sociodemographic and behavioural characteristics collected immediately prior to the pandemic, we used data from Natsal-3 to compare trends in differential uptake of interventions. Natsal-3 data provided the best comparison for these population-level sexual health interventions—with four key caveats. First, Natsal-3 data were collected ten years ago, and sexual behaviours and service provision have likely undergone secular changes in this timeframe. Second, Natsal-3 was a household-based probability sample, and there are different sampling biases between the surveys that weighting can only partially correct.^13^ Third, where we saw differences in associations, it was not possible to determine whether this was because of a change in the risk group, or a change in the reference group (or both). Likewise, where there was no difference between the surveys, this might still be an artefact caused by methodological differences. Finally, it was not possible to determine whether differences with Natsal-3 were pandemic-related or indicative of longer-term secular trends. Therefore, while the associations in the Natsal-COVID are strikingly similar to those seen in Natsal-3, these comparisons should be interpreted with caution.

### Conclusions and policy implications

Our study suggests differential access to key primary and secondary STI/HIV prevention interventions during the first year of the COVID-19 pandemic. However, the available evidence does not suggest substantial changes in inequalities in the patterns of uptake since 2010–12. While the pandemic might not have exacerbated inequalities in access to primary and secondary prevention, we did observe that large inequalities persist. These were typically among those at greatest STI/HIV risk, and there continues to be a need to reduce if not eradicate these. Future comparison with the fourth decennial probability survey (Natsal-4), which starts fieldwork in 2022, will be critical to continue to monitor inequalities and trends more broadly.

## Data Availability

An anonymised dataset will be deposited with the UK Data Archive.

## Contributors

The paper was conceived by ED, PS, JG, CHM, KRM, and NF. ED wrote the first draft, with further contributions from ED, PS, JG, AC, MW, JR, RBP, AJC, CT, CB, CO, SC, MU, CHM, KRM, and NF. Statistical analysis was done by ED. AC led on questionnaire design and data management was undertaken by JR and AC. PS and CHM are Principal Investigators (PIs) on Natsal and NF and KRM are PIs on Natsal-COVID. All authors contributed to data interpretation, reviewed successive drafts, and approved the final version of the manuscript.

## Acknowledgements

Natsal is a collaboration between University College London (UCL), the London School of Hygiene and Tropical Medicine (LSHTM), the University of Glasgow, Örebro University Hospital, and NatCen Social Research. The Natsal Resource, which is supported by a grant from the Wellcome Trust (212931/Z/18/Z), with contributions from the Economic and Social Research Council (ESRC) and National Institute for Health Research (NIHR), supports the Natsal-COVID study in addition to funding from the UCL Coronavirus Response Fund and the MRC/CSO Social and Public Health Sciences Unit (Core funding, MC_UU_00022/3; SPHSU18). We thank the study participants and Margaret Blake and Reuben Balfour (Ipsos MORI).

## Ethics approval

We obtained ethics approval from University of Glasgow MVLS College Ethics Committee (reference 20019174) and London School of Hygiene and Tropical Medicine Research Ethics committee (reference 22565).

## Data availability statement

An anonymised dataset will be deposited with the UK Data Archive.

