## Supplemental table for "How did the COVID-19 pandemic affect access to condoms, chlamydia and HIV testing, and cervical cancer screening at a population level in Britain? (Natsal-COVID)"

**Supplementary Material**

**Supplementary table 1**. Variations in reporting a chlamydia test among sexually-experienced women and men aged 18-44 years in the first year following the start of a national lockdown in Britain (23/03/2020) compared with Natsal-3 (2010-12)

|  | **Women (sexually-experienced)** | | | | | | | | | | | | | | |
| --- | --- | --- | --- | --- | --- | --- | --- | --- | --- | --- | --- | --- | --- | --- | --- |
|  | **Natsal COVID (fieldwork 2021)** | | | | | | | **Natsal-3 (fieldwork 2010-12)** | | | | | | |  |
|  | **Weighted %** | **95% CI** | **OR** | **95% CI** | **aOR ^*^** | **95% CI** | **Denominator ^†^ (unweighted, weighted)** | **Weighted %** | **95% CI** | **OR** | **95% CI** | **aOR ^*^** | **95% CI** | **Denominator ^†^ (unweighted, weighted)** | **Interaction terms between surveys p-value** |
| **All ages (18-44 years)** | 7.3% | [6.2%,8.5%] | - | - | - | - | 2145, 1824 | 25.1% | [23.7%,26.4%] | - | - | - | - | 5004, 3546 |  |
| **Age (years)** |  |  | **p<0.001** |  |  |  |  |  |  | **p<0.001** |  |  |  |  | **p=0.06** |
| 18-24 | 15.60% | [12.1%,20.0%] | 7.46 | (4.18 - 13.31) | - | - | 411, 328 | 54.5% | [51.5%,57.5%] | 11.97 | (9.42 - 15.22) | - | - | 1457, 833 |  |
| 25-29 | 10.70% | [8.1%,13.8%] | 4.79 | (2.69 - 8.54) | - | - | 545, 440 | 30.0% | [27.1%,33.0%] | 4.27 | (3.33 - 5.49) | - | - | 1357, 667 |  |
| 30-34 | 4.90% | [3.2%,7.5%] | 2.07 | (1.06 - 4.05) | - | - | 429, 355 | 16.5% | [14.2%,19.2%] | 1.98 | (1.50 - 2.61) | - | - | 1018, 645 |  |
| 35-44 | 2.40% | [1.5%,3.9%] | 1.00 |  | - | - | 760, 702 | 9.1% | [7.5%,11.0%] | 1.00 |  | - | - | 1172, 1402 |  |
| **Region** |  |  | **p=0.11** |  | **p=0.11** |  |  |  |  | **p<0.001** |  | **p<0.001** |  |  | **p=0.71** |
| England/Wales | 7.6% | [6.4%,9.0%] | 1.00 |  | 1.00 |  | 1955, 1649 | 25.7% | [24.3%,27.2%] | 1.00 |  | 1.00 |  | 4578, 3233 |  |
| Scotland | 4.1% | [1.9%,8.6%] | 0.52 | (0.23 - 1.17) | 0.50 | (0.22 - 1.16) | 190, 175 | 17.9% | [14.8%,21.5%] | 0.63 | (0.49 - 0.80) | 0.56 | (0.42 - 0.74) | 426, 313 |  |
| **Rurality** |  |  | **p=0.25** |  | **p=0.30** |  |  | - | - | - | - | - | - | - | - |
| Urban | 7.1% | [5.8%,8.7%] | 1.00 |  | 1.00 |  | 1508, 1281 | - | - | - | - | - | - | - | - |
| Rural | 5.1% | [3.0%,8.6%] | 0.71 | (0.39 - 1.28) | 0.71 | (0.38 - 1.36) | 254, 206 | - | - | - | - | - | - | - | - |
| **Ethnicity** |  |  | **p<0.001** |  | **p=0.001** |  |  |  |  | **p<0.001** |  | **p<0.001** |  |  | **p=0.25** |
| White ^1^ | 7.00% | [5.9%,8.3%] | 1.00 |  | 1.00 |  | 1839, 1498 | 25.5% | [24.1%,26.9%] | 1.00 |  | 1.00 |  | 4362, 3052 |  |
| Mixed, multiple, or other ^2^ | 16.60% | [8.4%,30.2%] | 2.65 | (1.19 - 5.90) | 2.42 | (1.06 - 5.55) | 82, 62 | 34.8% | [26.6%,43.9%] | 1.56 | (1.05 - 2.31) | 1.19 | (0.78 - 1.83) | 180, 116 |  |
| Asian or Asian British ^3^ | 1.80% | [0.5%,5.8%] | 0.24 | (0.07 - 0.83) | 0.19 | (0.05 - 0.71) | 141, 159 | 11.6% | [8.1%,16.3%] | 0.38 | (0.26 - 0.57) | 0.37 | (0.24 - 0.58) | 284, 237 |  |
| Black or Black British ^4^ | 19.30% | [10.9%,31.9%] | 3.19 | (1.59 - 6.40) | 2.25 | (1.12 - 4.53) | 63, 77 | 31.4% | [23.1%,41.0%] | 1.34 | (0.87 - 2.05) | 1.42 | (0.91 - 2.19) | 169, 134 |  |
| **Sexual identity** |  |  | **p=0.34** |  | **p=0.51** |  |  |  |  | **p=0.05** |  | **p=0.95** |  |  | **p=0.61** |
| Heterosexual/straight | 7.0% | [5.9%,8.3%] | 1.00 |  | 1.00 |  | 1845, 1728 | 24.9% | [23.5%,26.3%] | 1.00 |  | 1.00 |  | 4786, 3400 |  |
| Gay or lesbian | 16.6% | [8.4%,30.2%] | 0.61 | (0.18 - 2.06) | 0.46 | (0.12 - 1.79) | 66, 20 | 20.6% | [11.7%,33.5%] | 0.78 | (0.40 - 1.53) | 0.83 | (0.37 - 1.86) | 64, 47 |  |
| Bisexual | 1.8% | [0.5%,5.8%] | 1.43 | (0.85 - 2.40) | 1.07 | (0.62 - 1.84) | 187, 44 | 35.9% | [27.8%,45.0%] | 1.70 | (1.15 - 2.49) | 1.09 | (0.67 - 1.79) | 131, 83 |  |
| Other | - | - | - | - | - | - | 28, 11 ^**^ | - | - | - | - | - | - | 14, 10 ^**^ |  |
| **Social grade** |  |  | **p=0.16** |  | **p=0.51** |  |  | - | - | - | - | - | - | - | - |
| AB Higher and intermediate managerial/administrative/professional occupation | 7.20% | [5.1%,10.0%] | 1.00 |  | 1.00 |  | 500, 393 | - | - | - | - | - | - | - | - |
| C1 Supervisory, clerical and junior managerial/administrative/professional occupations/C2 Skilled manual occupations | 6.30% | [4.9%,8.1%] | 0.87 | (0.56 - 1.36) | 0.86 | (0.55 - 1.36) | 1042, 933 | - | - | - | - | - | - | - | - |
| D Semi-skilled and unskilled manual occupations/E On state benefit, unemployed and lowest grade occupations | 9.10% | [6.9%,11.9%] | 1.29 | (0.81 - 2.07) | 1.10 | (0.68 - 1.79) | 603, 498 | - | - | - | - | - | - | - | - |
| **Highest education qualification** |  |  | **p=0.52** |  | **p=0.97** |  |  |  |  | **p=0.001** |  | **p=0.93** |  |  | **p=0.53** |
| Degree | 6.70% | [5.3%,8.5%] | 1.00 |  | 1.00 |  | 1086, 930 | 21.4% | [19.1%,23.8%] | 1.00 |  | 1.00 |  | 1441, 1123 |  |
| Below degree | 7.70% | [6.1%,9.7%] | 1.16 | (0.81 - 1.67) | 0.96 | (0.66 - 1.39) | 955, 802 | 26.9% | [25.3%,28.7%] | 1.36 | (1.15 - 1.60) | 1.03 | (0.86 - 1.23) | 3239, 2221 |  |
| No qualifications | 9.30% | [5.0%,16.7%] | 1.33 | (0.70 - 2.94) | 0.95 | (0.42 - 2.15) | 104, 92 | 25.1% | [20.3%,30.5%] | 1.23 | (0.91 - 1.67) | 1.06 | (0.76 - 1.47) | 318, 199 |  |
| **Born outside the UK** |  |  | **p=0.56** |  | **p=0.54** |  |  | - | - | - | - | - | - | - | - |
| No | 7.20% | [6.0%,8.5%] | 1.00 |  | 1.00 |  | 1808, 1509 | - | - | - | - | - | - | - | - |
| Yes | 8.20% | [5.4%,12.1%] | 1.15 | (0.72 - 1.86) | 1.16 | (0.72 - 1.88) | 321, 299 | - | - | - | - | - | - | - | - |
| **Relationship status** |  |  | **p<0.001** |  | **p=0.001** |  |  |  |  | **p<0.001** |  | **p<0.001** |  |  | **p=0.40** |
| Married/steady and living together | 4.70% | [3.6%,6.1%] | 1.00 |  | 1.00 |  | 1277, 1104 | 15.7% | [14.4%,17.2%] | 1.00 |  | 1.00 |  | 2696, 2273 |  |
| Steady not living together | 11.50% | [7.9%,16.4%] | 2.63 | (1.61 - 4.32) | 1.91 | (1.14 - 3.18) | 255, 202 | 47.4% | [43.8%,51.1%] | 4.84 | (4.04 - 5.80) | 2.37 | (1.94 - 2.89) | 1016, 532 |  |
| Not in a steady relationship | 11.10% | [8.6%,14.2%] | 2.53 | (1.71 - 3.75) | 2.05 | (1.37 - 3.08) | 607, 512 | 37.4% | [34.3%,40.7%] | 3.21 | (2.70 - 3.82) | 2.09 | (1.73 - 2.53) | 1281, 735 |  |
| **Days drinking, past 7 days** |  |  | **p=0.05** |  | **p=0.04** |  |  | - | - | - | - | - | - | - | - |
| 0 days | 5.6% | [4.2%,7.3%] | 1.00 |  | 1.00 |  | 933, 805 | - | - | - | - | - | - | - | - |
| 1-2 days | 8.4% | [6.5%,10.8%] | 1.56 | (1.04 - 2.33) | 1.36 | (0.90 - 2.05) | 808, 678 | - | - | - | - | - | - | - | - |
| 3-4 days | 8.1% | [5.2%,12.6%] | 1.50 | (0.85 - 2.65) | 1.27 | (0.71 - 2.28) | 265, 228 | - | - | - | - | - | - | - | - |
| 5-7 days | 11.3% | [6.8%,18.3%] | 2.16 | (1.14 - 4.09) | 2.50 | (1.32 - 4.73) | 133, 107 | - | - | - | - | - | - | - | - |
| **Currently smoker** |  |  | **p=0.42** |  | **p=0.65** |  |  |  |  | **p<0.001** |  | **p<0.001** |  |  | **p=0.18** |
| No | 7.0% | [5.7%,8.4%] | 1.00 |  | 1.00 |  | 1647, 1403 | 22.1% | [20.6%,23.7%] | 1.00 |  | 1.00 |  | 3418, 2544 |  |
| Yes | 8.1% | [5.8%,11.3%] | 1.19 | (0.78 - 1.79) | 1.10 | (0.72 - 1.70) | 486, 411 | 32.5% | [29.9%,35.2%] | 1.69 | (1.46 - 1.97) | 1.49 | (1.28 - 1.75) | 1586, 1002 |  |
| **Importance of sexual health, past year** |  |  | **p<0.001** |  | **p<0.001** |  |  | - | - | - | - | - | - | - | - |
| Very important/somewhat important | 9.8% | [8.3%,11.7%] | 1.00 |  | 1.00 |  | 1372, 1166 | - | - | - | - | - | - | - | - |
| Not very important/not important | 2.3% | [1.4%,3.9%] | 0.22 | (0.12 - 0.39) | 0.24 | (0.14 - 0.43) | 605, 517 | - | - | - | - | - | - | - | - |
| This does not apply to me | 3.6% | [1.2%,10.6%] | 0.34 | (0.11 - 1.10) | 0.39 | (0.12 - 1.24) | 128, 108 | - | - | - | - | - | - | - | - |
| **Symptoms of depression (PHQ-2) ^5^** |  |  | **p=0.35** |  | **p=0.55** |  |  |  |  | **p=0.02** |  | **p=0.08** |  |  | **p=0.05** |
| No | 6.7% | [5.4%,8.3%] | 1.00 |  | 1.00 |  | 1300, 1119 | 24.6% | [23.2%,26.0%] | 1.00 |  | 1.00 |  | 4389, 3139 |  |
| Yes | 7.9% | [6.1%,10.2%] | 1.19 | (0.83 - 1.71) | 0.89 | (0.60 - 1.31) | 805, 671 | 29.4% | [25.7%,33.4%] | 1.28 | (1.05 - 1.55) | 1.22 | (0.98 - 1.52) | 605, 399 |  |
| **Symptoms of anxiety (GAD-2) ^5^** |  |  | **p=0.10** |  | **p=0.69** |  |  | - | - | - | - | - | - | - | - |
| No | 6.5% | [5.2%,8.1%] | 1.00 |  | 1.00 |  | 1228, 1075 | - | - | - | - | - | - | - | - |
| Yes | 8.5% | [6.7%,10.8%] | 1.34 | (0.94 - 1.90) | 1.08 | (0.74 - 1.57) | 899, 732 | - | - | - | - | - | - | - | - |
| **Total sexual partners, past year ^6^** |  |  | **p<0.001** |  | **p<0.001** |  |  |  |  | **p<0.001** |  | **p<0.001** |  |  | **p=0.31** |
| 0 partners | 3.3% | [1.9%,5.5%] | 1.00 |  | 1.00 |  | 472, 412 | 12.4% | [8.4%,17.9%] | 1.00 |  | 1.00 |  | 280, 184 |  |
| 1 partner | 6.4% | [5.2%,7.9%] | 2.02 | (1.12 - 3.63) | 2.21 | (1.19 - 4.10) | 1374, 1157 | 19.7% | [18.3%,21.1%] | 1.73 | (1.12 - 2.67) | 1.41 | (0.87 - 2.29) | 3687, 2758 |  |
| 2+ partners | 27.9% | [21.0%,36.0%] | 11.44 | (5.91 - 22.14) | 9.42 | (4.77 - 18.63) | 179, 146 | 54.2% | [50.4%,57.9%] | 8.32 | (5.19 - 13.34) | 4.69 | (2.78 - 7.91) | 998, 577 |  |
| **New sexual partners, past year ^6^** |  |  | **p<0.001** |  | **p<0.001** |  |  |  |  | **p<0.001** |  | **p<0.001** |  |  | **p=0.05** |
| 0 partners | 4.5% | [3.6%,5.7%] | 1.00 |  | 1.00 |  | 1695, 1443 | 17.7% | [16.4%,19.0%] | 1.00 |  | 1.00 |  | 3582, 2707 |  |
| 1 partner | 18.4% | [13.5%,24.7%] | 4.74 | (3.06 - 7.36) | 3.70 | (2.33 - 5.89) | 233, 196 | 43.1% | [39.1%,47.1%] | 3.52 | (2.93 - 4.24) | 2.30 | (1.90 - 2.80) | 854, 509 |  |
| 2+ partners | 36.8% | [26.3%,48.7%] | 12.23 | (7.10 - 21.07) | 9.54 | (5.48 - 16.59) | 91, 71 | 61.0% | [55.6%,66.0%] | 7.28 | (5.72 - 9.26) | 4.22 | (3.21 - 5.54) | 526, 299 |  |
| **Condom-less sex with a new partner on first occasion, past year ^6^** |  |  | **p<0.001** |  | **p<0.001** |  |  | - | - | - | - | - | - | - | - |
| None | 5.40% | [4.4%,6.5%] | 1.00 |  | 1.00 |  | 1845, 1555 | - | - | - | - | - | - | - | - |
| At least one | 26.10% | [19.8%,33.5%] | 6.23 | (4.11 - 9.44) | 5.05 | (3.27 - 7.79) | 191, 168 | - | - | - | - | - | - | - | - |
| **Previous same-sex experience, past 5 years ^7^** |  |  | **p=0.09** |  | **p=0.13** |  |  |  |  | **p<0.001** |  | **p=0.04** |  |  | **p=0.61** |
| No | 7.2% | [6.0%,8.5%] | 1.00 |  | 1.00 |  | 1977, 1740 | 24.4% | [23.0%,25.8%] | 1.00 |  | 1.00 |  | 4709, 3361 |  |
| Yes | 12.4% | [6.8%,21.5%] | 1.83 | (0.92 - 3.65) | 1.71 | (0.86 - 3.42) | 143, 65 | 36.9% | [30.9%,43.3%] | 1.81 | (1.37 - 2.40) | 1.39 | (1.01 - 1.92) | 294, 185 |  |
| **Used an STI-related service, past year** |  |  | **p<0.001** |  | **p<0.001** |  |  |  |  | **p<0.001** |  | **p<0.001** |  |  | **p=0.01** |
| No | 3.1% | [2.3%,4.1%] | 1.00 |  | 1.00 |  | 1912, 1621 | 19.5% | [18.3%,20.8%] | 1.00 |  | 1.00 |  | 4465, 3235 |  |
| Yes | 54.9% | [46.5%,62.9%] | 38.11 | (24.60 - 59.03) | 30.51 | (19.37 - 48.05) | 174, 146 | 83.1% | [78.8%,86.7%] | 20.23 | (15.17 - 26.98) | 13.05 | (9.57 - 17.81) | 532, 306 |  |
| **Unmet need for condoms, past year** |  |  | **p=0.07** |  | **p=0.57** |  |  | - | - | - | - | - | - | - | - |
| No | 7.30% | [6.2%,8.7%] | 1.00 |  | 1.00 |  | 1852, 1559 | - | - | - | - | - | - | - | - |
| Yes | 12.50% | [7.2%,20.9%] | 1.81 | (0.96 - 3.43) | 1.22 | (0.62 - 2.41) | 134, 116 | - | - | - | - | - | - | - | - |

|  | **Men (sexually-experienced)** | | | | | | | | | | | | | | |
| --- | --- | --- | --- | --- | --- | --- | --- | --- | --- | --- | --- | --- | --- | --- | --- |
|  | **Natsal COVID (fieldwork 2021)** | | | | | | | **Natsal-3 (fieldwork 2010-12)** | | | | | | |  |
|  | **Weighted %** | **95% CI** | **OR** | **95% CI** | **aOR ^*^** | **95% CI** | **Denominator ^†^ (unweighted, weighted)** | **Weighted %** | **95% CI** | **OR** | **95% CI** | **aOR ^*^** | **95% CI** | **Denominator ^†^ (unweighted, weighted)** | **Interaction terms between surveys p-value** |
| **All ages (18-44 years)** | 4.10% | [3.2%,5.3%] | - | - | - | - | 1666, 1870 | 15.10% | [13.9%,16.3%] | - | - | - | - | 3361, 3534 |  |
| **Age (years)** |  |  | **p=0.001** |  |  |  |  |  |  | **p<0.001** |  |  |  |  | **p=0.0070** |
| 18-24 | 7.00% | [4.4%,10.9%] | 3.75 | (1.90 - 7.42) | - | - | 358, 399 | 33.50% | [30.5%,36.7%] | 11.79 | (8.07 - 17.21) | - | - | 1134, 860 |  |
| 25-29 | 5.00% | [3.0%,8.3%] | 2.63 | (1.28 - 5.40) | - | - | 355, 370 | 19.40% | [16.7%,22.4%] | 5.62 | (3.79 - 8.33) | - | - | 823, 649 |  |
| 30-34 | 5.00% | [2.7%,9.1%] | 2.62 | (1.18 - 5.83) | - | - | 244, 287 | 9.50% | [7.2%,12.3%] | 2.44 | (1.55 - 3.86) | - | - | 621, 645 |  |
| 35-44 | 2.00% | [1.2%,3.1%] | 1.00 |  | - | - | 709, 815 | 4.10% | [2.9%,5.7%] | 1.00 |  | - | - | 783, 1380 |  |
| **Region** |  |  | **p=0.25** |  | **p=0.24** |  |  |  |  | **p=0.04** |  | **p=0.0206** |  |  | **p=0.6067** |
| England/Wales | 4.30% | [3.3%,5.6%] | 1.00 |  | 1.00 |  | 1526, 1702 | 15.50% | [14.2%,16.9%] | 1.00 |  | 1.00 |  | 3080, 3232 |  |
| Scotland | 2.10% | [0.6%,6.9%] | 0.47 | (0.13 - 1.70) | 0.46 | (0.12 - 1.68) | 140, 168 | 10.20% | [6.7%,15.1%] | 0.62 | (0.39 - 0.98) | 0.58 | (0.37 - 0.92) | 281, 302 |  |
| **Rurality** |  |  | **p=0.22** |  | **p=0.23** |  |  | - | - | - | - | - | - | - | - |
| Urban | 4.10% | [3.1%,5.5%] | 1.00 |  | 1.00 |  | 1245, 1397 | - | - | - | - | - | - | - | - |
| Rural | 1.90% | [0.6%,6.1%] | 0.46 | (0.14 - 1.57) | 0.47 | (0.14 - 1.61) | 134, 158 | - | - | - | - | - | - | - | - |
| **Ethnicity** |  |  | **p<0.001** |  | **p=0.001** |  |  |  |  | **p<0.001** |  | **p<0.001** |  |  | **p=0.45** |
| White ^1^ | 3.50% | [2.7%,4.7%] | 1.00 |  | 1.00 |  | 1360, 1509 | 15.40% | [14.1%,16.8%] | 1.00 |  | 1.00 |  | 2929, 2997 |  |
| Mixed, multiple, or other ^2^ | 17.10% | [7.6%,34.2%] | 5.63 | (2.14 - 14.83) | 4.66 | (1.77 - 12.21) | 61, 78 | 25.50% | [17.3%,35.8%] | 1.88 | (1.14 - 3.08) | 1.55 | (0.96 - 2.51) | 112, 112 |  |
| Asian or Asian British ^3^ | 1.40% | [0.4%,5.2%] | 0.39 | (0.10 - 1.55) | 0.35 | (0.08 - 1.43) | 146, 181 | 4.80% | [2.8%,8.1%] | 0.28 | (0.16 - 0.49) | 0.31 | (0.17 - 0.55) | 198, 286 |  |
| Black or Black British ^4^ | 9.30% | [4.0%,20.2%] | 2.79 | (1.07 - 7.24) | 2.26 | (0.87 - 5.84) | 65, 72 | 20.50% | [13.4%,30.1%] | 1.42 | (0.84 - 2.39) | 1.58 | (0.85 - 2.92) | 116, 133 |  |
| **Sexual identity** |  |  | **p<0.001** |  | **p<0.001** |  |  |  |  | **p<0.001** |  | **p<0.001** |  |  | **p=0.58** |
| Heterosexual/straight | 3.70% | [2.7%,4.9%] | 1.00 |  | 1.00 |  | 1431, 1768 | 14.50% | [13.3%,15.8%] | 1.00 |  | 1.00 |  | 3236, 3422 |  |
| Gay | 17.90% | [11.6%,26.6%] | 5.73 | (3.17 - 10.34) | 5.74 | (3.11 - 10.60) | 121, 49 | 42.10% | [29.7%,55.7%] | 4.29 | (2.46 - 7.48) | 4.54 | (2.24 - 9.18) | 82, 69 |  |
| Bisexual | 5.60% | [1.8%,16.4%] | 1.57 | (0.46 - 5.34) | 1.02 | (0.27 - 3.82) | 75, 22 | 16.90% | [7.4%,34.1%] | 1.20 | (0.47 - 3.06) | 0.90 | (0.35 - 2.30) | 32, 34 ^***^ |  |
| Other | - | - | - | - | - | - | 21, 12 ^**^ | - | - | - | - | - | - | 8, 5 ^**^ |  |
| **Social grade** |  |  | **p=0.01** |  | **p=0.01** |  |  | - | - | - | - | - | - | - | - |
| AB Higher and intermediate managerial/administrative/professional occupation | 6.40% | [4.4%,9.1%] | 1.00 |  | 1,00 |  | 580, 433 | - | - | - | - | - | - | - | - |
| C1 Supervisory, clerical and junior managerial/administrative/professional occupations/C2 Skilled manual occupations | 2.60% | [1.6%,4.1%] | 0.39 | (0.21 - 0.72) | 0.39 | (0.21 - 0.73) | 691, 1010 | - | - | - | - | - | - | - | - |
| D Semi-skilled and unskilled manual occupations/E On state benefit, unemployed and lowest grade occupations | 5.40% | [3.3%,8.9%] | 0.84 | (0.44 - 1.63) | 0.75 | (0.39 - 1.43) | 395, 427 | - | - | - | - | - | - | - | - |
| **Highest education qualification** |  |  | **p<0.001** |  | **p=0.005** |  |  |  |  | **p<0.001** |  | **p=0.70** |  |  | **p=0.04** |
| Degree | 3.10% | [2.1%,4.5%] | 1.00 |  | 1.00 |  | 810, 845 | 11.40% | [9.5%,13.7%] | 1.00 |  | 1.00 |  | 906, 1057 |  |
| Below degree | 3.80% | [2.6%,5.5%] | 1.23 | (0.72 - 2.13) | 1.10 | (0.64 - 1.89) | 750, 905 | 17.10% | [15.5%,18.8%] | 1.60 | (1.26 - 2.02) | 1.11 | (0.87 - 1.42) | 2277, 2273 |  |
| No qualifications | 13.70% | [6.9%,25.2%] | 4.97 | (2.13 - 11.59) | 3.93 | (1.70 - 9.11) | 106, 120 | 11.30% | [7.1%,17.4%] | 0.98 | (0.57 - 1.70) | 1.03 | (0.55 - 1.92) | 173, 200 |  |
| **Born outside the UK** |  |  | **p=0.38** |  | **p=0.47** |  |  | - | - | - | - | - | - | - | - |
| No | 4.30% | [3.3%,5.6%] | 1.00 |  | 1.00 |  | 1491, 1681 | - | - | - | - | - | - | - | - |
| Yes | 2.70% | [1.0%,7.3%] | 0.62 | (0.21 - 1.81) | 0.67 | (0.23 - 1.97) | 160, 174 | - | - | - | - | - | - | - | - |
| **Relationship status** |  |  | **p=0.54** |  | **p=0.45** |  |  |  |  | **p<0.001** |  | **p<0.001** |  |  | **p<0.001** |
| Married/steady and living together | 4.00% | [2.8%,5.7%] | 1.00 |  | 1.00 |  | 972, 1099 | 7.00% | [5.8%,8.3%] | 1.00 |  | 1.00 |  | 1622, 2153 |  |
| Steady not living together | 2.80% | [1.0%,7.6%] | 0.70 | (0.23 - 2.10) | 0.47 | (0.14 - 1.55) | 153, 174 | 30.40% | [26.7%,34.5%] | 5.84 | (4.46 - 7.65) | 2.58 | (1.87 - 3.56) | 676, 531 |  |
| Not in a steady relationship | 4.90% | [3.3%,7.0%] | 1.23 | (0.71 - 2.12) | 0.97 | (0.54 - 1.74) | 526, 577 | 25.90% | [23.1%,29.0%] | 4.68 | (3.64 - 6.01) | 2.30 | (1.72 - 3.07) | 1055, 842 |  |
| **Days drinking, past 7 days** |  |  | **p=0.03** |  | **p=0.03** |  |  | - | - | - | - | - | - | - | - |
| 0 days | 3.90% | [2.3%,6.6%] | 1.00 |  | 1.00 |  | 505, 559 | - | - | - | - | - | - | - | - |
| 1-2 days | 3.30% | [2.1%,5.1%] | 0.83 | (0.40 - 1.71) | 0.79 | (0.38 - 1.63) | 667, 780 | - | - | - | - | - | - | - | - |
| 3-4 days | 7.20% | [4.8%,10.8%] | 1.92 | (0.95 - 3.89) | 1.91 | (0.94 - 3.89) | 326, 350 | - | - | - | - | - | - | - | - |
| 5-7 days | 2.50% | [1.0%,6.2%] | 0.63 | (0.21 - 1.89) | 0.64 | (0.21 - 1.92) | 162, 174 | - | - | - | - | - | - | - | - |
| **Currently smoker** |  |  | **p<0.001** |  | **p<0.001** |  |  |  |  | **p<0.001** |  | **p<0.001** |  |  | **p=0.04** |
| No | 2.30% | [1.6%,3.2%] | 1.00 |  | 1.00 |  | 1135, 1255 | 12.40% | [11.1%,13.9%] | 1.00 |  | 1.00 |  | 2181, 2388 |  |
| Yes | 8.00% | [5.7%,11.3%] | 3.77 | (2.24 - 6.34) | 3.39 | (1.99 - 5.78) | 516, 599 | 20.60% | [18.1%,23.3%] | 1.83 | (1.49 - 2.25) | 1.64 | (1.32 - 2.02) | 1180, 1146 |  |
| **Importance of sexual health, past year** |  |  | **p=0.002** |  | **p=0.002** |  |  | - | - | - | - | - | - | - | - |
| Very important/somewhat important | 5.40% | [4.1%,7.2%] | 1.00 |  | 1.00 |  | 1037, 1184 | - | - | - | - | - | - | - | - |
| Not very important/not important | 2.10% | [1.2%,3.7%] | 0.37 | (0.19 - 0.72) | 0.36 | (0.19 - 0.70) | 494, 541 | - | - | - | - | - | - | - | - |
| This does not apply to me | 0.60% | [0.1%,4.5%] | 0.11 | (0.02 - 0.84) | 0.14 | (0.02 - 1.05) | 106, 111 | - | - | - | - | - | - | - | - |
| **Symptoms of depression (PHQ-2) ^5^** |  |  | **p=0.06** |  | **p=0.22** |  |  |  |  | **p=0.06** |  | **p=0.17** |  |  | **p=0.82** |
| No | 3.20% | [2.2%,4.5%] | 1.00 |  | 1.00 |  | 1004, 1127 | 14.70% | [13.4%,16.0%] | 1.00 |  | 1.00 |  | 3012, 3196 |  |
| Yes | 5.20% | [3.5%,7.7%] | 1.69 | (0.97 - 2.94) | 1.44 | (0.81 - 2.56) | 626, 706 | 18.90% | [14.8%,23.8%] | 1.35 | (0.98 - 1.85) | 1.28 | (0.90 - 1.82) | 343, 332 |  |
| **Symptoms of anxiety (GAD-2) ^5^** |  |  | **p=0.004** |  | **p=0.02** |  |  | - | - | - | - | - | - | - | - |
| No | 2.90% | [2.0%,4.1%] | 1.00 |  | 1.00 |  | 1066, 1193 | - | - | - | - | - | - | - | - |
| Yes | 6.20% | [4.3%,8.9%] | 2.23 | (1.30 - 3.84) | 1.94 | (1.11 - 3.38) | 570, 646 | - | - | - | - | - | - | - | - |
| **Total sexual partners, past year ^6^** |  |  | **p<0.001** |  | **p<0.001** |  |  |  |  | **p<0.001** |  | **p<0.001** |  |  | **p=0.004** |
| 0 partners | 2.60% | [1.5%,4.5%] | 1.00 |  | 1.00 |  | 412, 462 | 1.20% | [0.4%,3.8%] | 1.00 |  | 1.00 |  | 180, 168 |  |
| 1 partner | 2.00% | [1.3%,3.3%] | 0.78 | (0.36 - 1.65) | 0.88 | (0.41 - 1.90) | 923, 1056 | 9.80% | [8.6%,11.1%] | 9.08 | (2.71 - 30.44) | 11.24 | (3.34 - 37.83) | 2207, 2545 |  |
| 2+ partners | 15.60% | [10.8%,22.1%] | 6.93 | (3.38 - 14.18) | 6.60 | (3.22 - 13.55) | 223, 238 | 34.60% | [31.1%,38.3%] | 44.23 | (13.12 - 149.21) | 35.10 | (10.38 - 118.69) | 945, 795 |  |
| **New sexual partners, past year ^6^** |  |  | **p<0.001** |  | **p<0.001** |  |  |  |  | **p<0.001** |  | **p<0.001** |  |  | **p=0.18** |
| 0 partners | 1.70% | [1.1%,2.7%] | 1.00 |  | 1.00 |  | 1180, 1336 | 8.20% | [7.1%,9.5%] | 1.00 |  | 1.00 |  | 2059, 2462 |  |
| 1 partner | 7.40% | [4.6%,11.6%] | 4.54 | (2.31 - 8.92) | 4.09 | (2.06 - 8.09) | 226, 257 | 24.90% | [21.5%,28.7%] | 3.72 | (2.86 - 4.82) | 2.13 | (1.62 - 2.80) | 695, 576 |  |
| 2+ partners | 18.20% | [11.7%,27.2%] | 12.65 | (6.34 - 25.23) | 10.70 | (5.31 - 21.59) | 146, 155 | 39.00% | [34.3%,43.9%] | 7.16 | (5.51 - 9.29) | 4.31 | (3.23 - 5.77) | 573, 464 |  |
| **Condom-less sex with a new partner on first occasion, past year ^6^** |  |  | **p<0.001** |  | **p<0.001** |  |  | - | - | - | - | - | - | - | - |
| None | 2.20% | [1.5%,3.1%] | 1.00 |  | 1.00 |  | 1288, 1446 | - | - | - | - | - | - | - | - |
| At least one | 13.20% | [9.1%,18.6%] | 6.89 | (3.91 - 12.15) | 6.14 | (3.46 - 10.87) | 260, 297 | - | - | - | - | - | - | - | - |
| **Previous same-sex experience, past 5 years ^7^** |  |  | **p<0.001** |  | **p<0.001** |  |  |  |  | **p<0.001** |  | **p<0.001** |  |  | **p=0.04** |
| No | 3.00% | [2.2%,4.2%] | 1.00 |  | 1.00 |  | 1452, 1723 | 14.40% | [13.2%,15.7%] | 1.00 |  | 1.00 |  | 3231, 3422 |  |
| Yes | 17.30% | [11.5%,25.3%] | 6.79 | (3.77 - 12.24) | 6.43 | (3.51 - 11.78) | 183, 115 | 35.10% | [26.2%,45.1%] | 3.21 | (2.09 - 4.94) | 3.01 | (1.76 - 5.14) | 130, 112 |  |
| **Used an STI-related service, past year** |  |  | **p<0.001** |  | **p<0.001** |  |  |  |  | **p<0.001** |  | **p<0.001** |  |  | **p<0.001** |
| No | 2.40% | [1.6%,3.5%] | 1.00 |  | 1.00 |  | 1457, 1648 | 9.90% | [8.9%,11.0%] | 1.00 |  | 1.00 |  | 3044, 3283 |  |
| Yes | 23.10% | [16.9%,30.7%] | 12.43 | (7.06 - 21.88) | 10.81 | (5.78 - 20.23) | 163, 163 | 84.0% | [79.1%,87.9%] | 47.70 | (33.62 - 67.69) | 36.84 | (23.95 - 56.67) | 314, 247 |  |
| **Unmet need for condoms, past year** |  |  | **p<0.001** |  | **p<0.001** |  |  | - | - | - | - | - | - | - | - |
| No | 2.60% | [1.8%,3.6%] | 1.00 |  | 1.00 |  | 1262, 1406 | - | - | - | - | - | - | - | - |
| Yes | 14.20% | [9.8%,20.1%] | 6.30 | (3.61 - 11.00) | 5.23 | (2.88 - 9.51) | 238, 270 | - | - | - | - | - | - | - | - |

CI=confidence intervals. OR=odds ratio. aOR=age-adjusted odds ratio. PHQ-2=Patient Health Questionnaire (2 item). GAD-2=Generalized anxiety disorder (2 item)

^*^ Age adjusted

^†^ Men or women aged 18-44 who were sexually-experienced. Trans men and trans women are included in data for men and women, respectively. 18 women and 24 men in Natsal-COVID responded 'prefer not to say' to questions about chlamydia testing. 120 women and 66 men in Natsal-3 did not answer the question. These individuals are excluded from the denominator.

^**^ Unweighted denominator <30. Results not shown due to small denominator

^***^Unweighted denominator <50. Results should be interpreted with caution due to small denominator.

^1^ White includes all those who identify as White English, Welsh, Scottish, Northern Irish, British, Irish, Gypsy or Irish Traveller, or from any other White background.

^2^ Mixed ethnicity includes those who identify as White and Black African, White and Black Caribbean, White and Asian or any other mixed or multiple ethnic background.

^3^ Asian includes those who identify as Indian, Pakistani, Bangladeshi, Chinese or from any other Asian background

^4^ Black includes those who identify as African, Caribbean, or from any other Black background.

^5^ Participants were classified as having symptoms of depression or anxiety if they scored three or more on the patient health questionnaire two item (PHQ-2) or generalised anxiety disorder two item (GAD-2) scales

^6^ Includes both opposite-sex and same-sex partners

^7^ Same-sex experience defined as oral/anal/vaginal sex

All percentages are weighted. These are row percentages which describe reported chlamydia testing in the past year within certain subgroups.

**Supplementary table 2.** Variations in reporting an HIV test among sexually-experienced women and men aged 18-44 years in the first year following the start of a national lockdown in Britain (23/03/2020) compared with Natsal-3 (2010-12)

|  | **Women (sexually-experienced)** | | | | | | | | | | | | | | |
| --- | --- | --- | --- | --- | --- | --- | --- | --- | --- | --- | --- | --- | --- | --- | --- |
|  | **Natsal COVID (fieldwork 2021)** | | | | | | | **Natsal-3 (fieldwork 2010-12)** | | | | | | |  |
|  | **Weighted %** | **95% CI** | **OR** | **95% CI** | **aOR ^*^** | **95% CI** | **Denominator ^†^ (unweighted, weighted)** | **Weighted %** | **95% CI** | **OR** | **95% CI** | **aOR ^*^** | **95% CI** | **Denominator ^†^ (unweighted, weighted)** | **Interaction terms between surveys p-value** |
| **All ages (18-44 years)** | 8.6% | [7.4%,10.0%] | - | - | - | - | 2148, 1827 | 10.4% | [9.5%,11.4%] | - | - | - | - | 4701, 3331 |  |
| **Age (years)** |  |  | **p<0.001** |  |  |  |  |  |  | **p<0.001** |  |  |  |  | **p=0.92** |
| 18-24 | 12.2% | [9.0%,16.4%] | 2.85 | (1.71 - 4.75) | - | - | 411, 327 | 14.9% | [12.8%,17.3%] | 3.11 | (2.22 - 4.35) | - | - | 1367, 783 |  |
| 25-29 | 11.8% | [9.0%,15.3%] | 2.76 | (1.70 - 4.47) | - | - | 545, 440 | 13.7% | [11.5%,16.1%] | 2.81 | (1.99 - 3.96) | - | - | 1269, 622 |  |
| 30-34 | 9.1% | [6.6%,12.4%] | 2.06 | (1.23 - 3.43) | - | - | 430, 356 | 12.3% | [10.3%,14.6%] | 2.48 | (1.74 - 3.53) | - | - | 970, 614 |  |
| 35-44 | 4.6% | [3.2%,6.6%] | 1.00 |  | - | - | 762, 703 | 5.3% | [4.1%,7.0%] | 1.00 |  | - | - | 1095, 1312 |  |
| **Region** |  |  | **p=0.01** |  | **p=0.008** |  |  |  |  | **p=0.02** |  | **p=0.02** |  |  | **p=0.29** |
| England/Wales | 9.1% | [7.8%,10.7%] | 1.00 |  | 1.00 |  | 1957, 1650 | 10.8% | [9.8%,11.8%] | 1.00 |  | 1.00 |  | 4307, 3036 |  |
| Scotland | 3.7% | [1.8%,7.2%] | 0.38 | (0.18 - 0.79) | 0.37 | (0.18 - 0.77) | 191, 176 | 6.7% | [4.6%,9.7%] | 0.59 | (0.39 - 0.90) | 0.59 | (0.38 - 0.90) | 394, 294 |  |
| **Rurality** |  |  | **p=0.10** |  | **p=0.13** |  |  | - | - | - | - | - | - | - | - |
| Urban | 8.4% | [7.0%,10.2%] | 1.00 |  | 1.00 |  | 1511, 1283 | - | - | - | - | - | - | - | - |
| Rural | 5.3% | [3.1%,8.9%] | 0.61 | (0.34 - 1.10) | 0.62 | (0.34 - 1.14) | 255, 207 | - | - | - | - | - | - | - | - |
| **Ethnicity** |  |  | **p<0.001** |  | **p<0.001** |  |  |  |  | **p<0.001** |  | **p<0.001** |  |  | **p=0.11** |
| White ^1^ | 7.4% | [6.2%,8.7%] | 1.00 |  | 1.00 |  | 1840, 1498 | 9.9% | [9.0%,10.9%] | 1.00 |  | 1.00 |  | 4076, 2850 |  |
| Mixed, multiple, or other ^2^ | 26.4% | [15.1%,41.8%] | 4.50 | (2.19 - 9.25) | 4.28 | (2.01 - 9.12) | 83, 63 | 15.0% | [10.0%,22.1%] | 1.61 | (0.99 - 2.63) | 1.39 | (0.84 - 2.33) | 173, 111 |  |
| Asian or Asian British ^3^ | 5.5% | [2.8%,10.9%] | 0.74 | (0.35 - 1.57) | 0.68 | (0.31 - 1.48) | 142, 159 | 7.2% | [4.4%,11.8%] | 0.71 | (0.41 - 1.23) | 0.74 | (0.42 - 1.29) | 277, 235 |  |
| Black or Black British ^4^ | 23.6% | [14.2%,36.6%] | 3.88 | (2.02 - 7.46) | 3.15 | (1.64 - 6.02) | 63, 77 | 23.1% | [16.2%,32.0%] | 2.74 | (1.73 - 4.33) | 2.80 | (1.80 - 4.33) | 166, 128 |  |
| **Sexual identity** |  |  | **p=0.65** |  | **p=0.39** |  |  |  |  | **p=0.44** |  | **p=0.79** |  |  | **p=0.21** |
| Heterosexual/straight | 8.8% | [7.5%,10.3%] | 1.00 |  | 1.00 |  | 1848, 1730 | 10.3% | [9.3%,11.3%] | 1.00 |  | 1.00 |  | 4494, 3191 |  |
| Gay or lesbian | 5.5% | [1.9%,14.6%] | 0.60 | (0.20 - 1.80) | 0.53 | (0.17 - 1.65) | 65, 19 | 12.3% | [6.0%,23.4%] | 1.23 | (0.56 - 2.70) | 1.29 | (0.59 - 2.80) | 60, 45 |  |
| Bisexual | 8.2% | [4.9%,13.4%] | 0.93 | (0.52 - 1.64) | 0.77 | (0.43 - 1.37) | 187, 44 | 14.8% | [9.4%,22.5%] | 1.52 | (0.90 - 2.56) | 1.24 | (0.72 - 2.14) | 125, 80 |  |
| Other | - |  | - | - | - | - | 29, 12 ^**^ | - |  | - | - | - | - | 14, 10 ^**^ |  |
| **Social grade** |  |  | **p=0.27** |  | **p=0.48** |  |  | - | - | - | - | - | - | - | - |
| AB Higher and intermediate managerial/administrative/professional occupation | 9.4% | [7.0%,12.5%] | 1.00 |  | 1.00 |  | 500, 393 | - | - | - | - | - | - | - | - |
| C1 Supervisory, clerical and junior managerial/administrative/professional occupations/C2 Skilled manual occupations | 7.5% | [5.9%,9.6%] | 0.79 | (0.52 - 1.20) | 0.80 | (0.53 - 1.22) | 1044, 934 | - | - | - | - | - | - | - | - |
| D Semi-skilled and unskilled manual occupations/E On state benefit, unemployed and lowest grade occupations | 10.0% | [7.6%,12.9%] | 1.07 | (0.69 - 1.66) | 0.98 | (0.62 - 1.52) | 604, 499 | - | - | - | - | - | - | - | - |
| **Highest education qualification** |  |  | **p=0.29** |  | **p=0.09** |  |  |  |  | **p=0.58** |  | **p=0.09** |  |  | **p=0.67** |
| Degree | 9.60% | [7.8%,11.8%] | 1.00 |  | 1.00 |  | 1088, 931 | 11.1% | [9.5%,13.0%] | 1.00 |  | 1.00 |  | 1372, 1066 |  |
| Below degree | 7.40% | [5.8%,9.4%] | 0.76 | (0.53 - 1.07) | 0.68 | (0.48 - 0.96) | 955, 804 | 10.0% | [8.9%,11.2%] | 0.89 | (0.72 - 1.11) | 0.78 | (0.63 - 0.98) | 3021, 2074 |  |
| No qualification | 8.80% | [4.5%,16.3%] | 0.91 | (0.43 - 1.91) | 0.70 | (0.32 - 1.55) | 105, 92 | 10.5% | [7.3%,14.8%] | 0.94 | (0.61 - 1.45) | 0.88 | (0.57 - 1.36) | 302, 186 |  |
| **Born outside the UK** |  |  | **p=0.13** |  | **p=0.14** |  |  | - | - | - | - | - | - | - | - |
| No | 8.1% | [6.9%,9.6%] | 1.00 |  | 1.00 |  | 1908, 1509 | - | - | - | - | - | - | - | - |
| Yes | 11.1% | [7.7%,15.9%] | 1.41 | (0.90 - 2.22) | 1.41 | (0.90 - 2.22) | 323, 301 | - | - | - | - | - | - | - | - |
| **Relationship status** |  |  | **p=0.003** |  | **p= 0.02** |  |  |  |  | **p=0.003** |  | **p=0.69** |  |  | **p=0.17** |
| Married/steady and living together | 6.80% | [5.5%,8.5%] | 1.00 |  | 1.00 |  | 1277, 1105 | 9.2% | [8.1%,10.5%] | 1.00 |  | 1.00 |  | 2518, 2132 |  |
| Steady not living together | 8.20% | [5.3%,12.6%] | 1.22 | (0.72 - 2.07) | 1.01 | (0.58 - 1.73) | 254, 201 | 12.8% | [10.5%,15.5%] | 1.44 | (1.11 - 1.88) | 0.93 | (0.70 - 1.24) | 951, 498 |  |
| Not in a steady relationship | 12.30% | [9.5%,15.8%] | 1.91 | (1.32 - 2.76) | 1.69 | (1.15 - 2.48) | 611, 515 | 12.5% | [10.5%,14.9%] | 1.41 | (1.11 - 1.79) | 1.06 | (0.83 - 1.35) | 1206, 687 |  |
| **Days drinking, past 7 days** |  |  | **p=0.12** |  | **p=0.13** |  |  | - | - | - | - | - | - | - | - |
| 0 days | 7.1% | [5.4%,9.1%] | 1.00 |  | 1.00 |  | 935, 807 | - | - | - | - | - | - | - | - |
| 1-2 days | 9.8% | [7.7%,12.4%] | 1.43 | (0.97 - 2.10) | 1.31 | (0.89 - 1.93) | 810, 680 | - | - | - | - | - | - | - | - |
| 3-4 days | 8.8% | [5.7%,13.3%] | 1.26 | (0.73 - 2.18) | 1.14 | (0.67 - 1.95) | 265, 227 | - | - | - | - | - | - | - | - |
| 5-7 days | 12.7% | [7.8%,20.1%] | 1.93 | (1.05 - 3.54) | 2.04 | (1.10 - 3.78) | 132, 107 | - | - | - | - | - | - | - | - |
| **Currently smoker** |  |  | **p=0.03** |  | **p=0.05** |  |  |  |  | **p=0.96** |  | **p=0.46** |  |  | **p=0.04** |
| No | 7.7% | [6.5%,9.3%] | 1.00 |  | 1.00 |  | 1652, 1408 | 10.4% | [9.3%,11.6%] | 1.00 |  | 1.00 |  | 3235, 2407 |  |
| Yes | 11.5% | [8.5%,15.4%] | 1.55 | (1.05 - 2.28) | 1.49 | (1.00 - 2.21) | 483, 408 | 10.5% | [8.9%,12.3%] | 1.01 | (0.80 - 1.26) | 0.92 | (0.73 - 1.15) | 1466, 923 |  |
| **Importance of sexual health, past year** |  |  | **p<0.001** |  | **p<0.001** |  |  | - | - | - | - | - | - | - | - |
| Very important/somewhat important | 10.9% | [9.1%,12.8%] | 1.00 |  | 1.00 |  | 1370, 1164 | - | - | - | - | - | - | - | - |
| Not very important/not important | 4.5% | [3.1%,6.7%] | 0.39 | (0.25 - 0.61) | 0.42 | (0.27 - 0.67) | 606, 517 | - | - | - | - | - | - | - | - |
| This does not apply to me | 4.7% | [1.8%,11.8%] | 0.40 | (0.15 - 1.12) | 0.43 | (0.16 - 1.20) | 130, 109 | - | - | - | - | - | - | - | - |
| **Symptoms of depression (PHQ-2) ^5^** |  |  | **p=0.55** |  | **p=0.75** |  |  |  |  | **p=0.81** |  | **p=0.89** |  |  | **p=0.71** |
| No | 8.1% | [6.6%,9.9%] | 1.00 |  | 1.00 |  | 1305, 1124 | 10.3% | [9.4%,11.4%] | 1.00 |  | 1.00 |  | 4132, 2945 |  |
| Yes | 9.0% | [6.9%,11.5%] | 1.11 | (0.78 - 1.59) | 0.94 | (0.66 - 1.35) | 803, 668 | 10.7% | [8.2%,13.9%] | 1.04 | (0.76 - 1.42) | 1.02 | (0.75 - 1.40) | 559, 376 |  |
| **Symptoms of anxiety (GAD-2) ^5^** |  |  | **p=0.24** |  | **p=0.68** |  |  | - | - | - | - | - | - | - | - |
| No | 7.9% | [6.4%,9.8%] | 1.00 |  | 1.00 |  | 1231, 1077 | - | - | - | - | - | - | - | - |
| Yes | 9.5% | [7.6%,12.0%] | 1.23 | (0.87 - 1.72) | 1.08 | (0.76 - 1.52) | 899, 731 | - | - | - | - | - | - | - | - |
| **Total sexual partners, past year ^6^** |  |  | **p<0.001** |  | **p<0.001** |  |  |  |  | **p<0.001** |  | **p<0.001** |  |  | **p<0.001** |
| 0 partners | 7.6% | [5.1%,11.2%] | 1.00 |  | 1.00 |  | 473, 413 | 4.6% | [2.3%,8.7%] | 1.00 |  | 1.00 |  | 266, 175 |  |
| 1 partner | 6.6% | [5.4%,8.1%] | 0.86 | (0.53 - 1.38) | 0.87 | (0.54 - 1.42) | 1376, 1159 | 9.2% | [8.2%,10.2%] | 2.11 | (1.04 - 4.26) | 1.92 | (0.95 - 3.89) | 3447, 2573 |  |
| 2+ partners | 32.0% | [24.6%,40.4%] | 5.73 | (3.27 - 10.05) | 5.03 | (2.87 - 8.80) | 179, 146 | 19.1% | [16.3%,22.3%] | 4.95 | (2.36 - 10.37) | 3.57 | (1.71 - 7.46) | 926, 533 |  |
| **New sexual partners, past year ^6^** |  |  | **p<0.001** |  | **p<0.001** |  |  |  |  | **p<0.001** |  | **p<0.001** |  |  | **p=0.001** |
| 0 partners | 6.6% | [5.4%,8.1%] | 1.00 |  | 1.00 |  | 1698, 1446 | 8.5% | [7.6%,9.6%] | 1.00 |  | 1.00 |  | 3348, 2521 |  |
| 1 partner | 16.5% | [11.9%,22.3%] | 2.79 | (1.81 - 4.30) | 2.43 | (1.57 - 3.74) | 233, 195 | 13.8% | [11.3%,16.8%] | 1.72 | (1.31 - 2.26) | 1.38 | (1.05 - 1.83) | 794, 475 |  |
| 2+ partners | 37.5% | [26.7%,49.6%] | 8.48 | (4.94 - 14.57) | 7.29 | (4.22 - 12.61) | 91, 71 | 22.9% | [18.7%,27.6%] | 3.18 | (2.39 - 4.23) | 2.36 | (1.75 - 3.18) | 494, 282 |  |
| **Condom-less sex with a new partner on first occasion, past year ^6^** |  |  | **p<0.001** |  | **p<0.001** |  |  | - | - | - | - | - | - | - | - |
| None | 7.3% | [6.1%,8.8%] | 1.00 |  | 1.00 |  | 1848, 1558 | - | - | - | - | - | - | - | - |
| At least one | 23.8% | [17.8%,31.1%] | 3.96 | (2.61 - 6.01) | 3.44 | (2.26 - 5.23) | 190, 167 | - | - | - | - | - | - | - | - |
| **Previous same-sex experience, past 5 years ^7^** |  |  | **p=0.005** |  | **p=0.009** |  |  |  |  | **p=0.008** |  | **p=0.07** |  |  | **p=0.20** |
| No | 8.3% | [7.0%,9.7%] | 1.00 |  | 1.00 |  | 1979, 1742 | 10.1% | [9.2%,11.2%] | 1.00 |  | 1.00 |  | 4428, 3154 |  |
| Yes | 17.9% | [10.8%,28.1%] | 2.41 | (1.31 - 4.45) | 2.27 | (1.23 - 4.19) | 143, 65 | 15.5% | [11.5%,20.5%] | 1.63 | (1.13 - 2.33) | 1.41 | (0.98 - 2.03) | 271, 174 |  |
| **Pregnant in past year or currently pregnant** |  |  | **p<0.001** |  | **p<0.001** |  |  |  |  | **p<0.001** |  | **p<0.001** |  |  | **p=0.01** |
| No | 6.9% | [5.6%,8.4%] | 1.00 |  | 1.00 |  | 1658, 1393 | 6.4% | [5.6%,7.3%] | 1.00 |  | 1.00 |  | 3878, 2815 |  |
| Yes | 23.3% | [18.0%,29.5%] | 4.11 | (2.78 - 6.06) | 3.94 | (2.65 - 5.86) | 221, 191 | 33.8% | [30.2%,37.7%] | 7.50 | (6.04 - 9.31) | 7.06 | (5.65 - 8.81) | 798, 494 |  |
| **Used an STI-related service, past year** |  |  | **p<0.001** |  | **p<0.001** |  |  |  |  | **p<0.001** |  | **p<0.001** |  |  | **p=0.02** |
| No | 4.9% | [3.9%,6.0%] | 1.00 |  | 1.00 |  | 1917, 1625 | 6.9% | [6.1%,7.7%] | 1.00 |  | 1.00 |  | 4151, 3003 |  |
| Yes | 51.6% | [43.3%,59.7%] | 20.87 | (13.95 - 31.23) | 20.03 | (13.03 - 30.78) | 173, 145 | 47.4% | [42.5%,52.3%] | 12.21 | (9.67 - 15.40) | 10.76 | (8.40 - 13.79) | 512, 297 |  |
| **Unmet need for condoms, past year** |  |  | **p<0.001** |  | **p<0.001** |  |  | - | - | - | - | - | - | - | - |
| No | 8.00% | [6.8%,9.5%] | 1.00 |  | 1.00 |  | 1856, 1563 | - | - | - | - | - | - | - | - |
| Yes | 19.10% | [12.5%,28.1%] | 2.70 | (1.58 - 4.62) | 2.16 | (1.22 - 3.85) | 133, 115 | - | - | - | - | - | - | - | - |

|  | **Men (sexually-experienced)** | | | | | | | | | | | | | | |
| --- | --- | --- | --- | --- | --- | --- | --- | --- | --- | --- | --- | --- | --- | --- | --- |
|  | **Natsal COVID (fieldwork 2021)** | | | | | | | **Natsal-3 (fieldwork 2010-12)** | | | | | | |  |
|  | **Weighted %** | **95% CI** | **OR** | **95% CI** | **aOR ^*^** | **95% CI** | **Denominator ^†^ (unweighted, weighted)** | **Weighted %** | **95% CI** | **OR** | **95% CI** | **aOR ^*^** | **95% CI** | **Denominator ^†^ (unweighted, weighted)** | **Interaction terms between surveys p-value** |
| **All ages (18-44 years)** | 6.5% | [5.4%,7.9%] | - | - | - | - | 1668, 1868 | 6.00% | [5.1%,7.0%] | - | - | - | - | 3198, 3366 |  |
| **Age (years)** |  |  | **p=0.005** |  |  |  |  |  |  | **p=0.002** |  |  |  |  | **p=0.49** |
| 18-24 | 8.6% | [5.9%,12.4%] | 2.28 | (1.31 - 3.97) | - | - | 359, 395 | 6.40% | [5.0%,8.2%] | 1.71 | (1.06 - 2.77) | - | - | 1082, 820 |  |
| 25-29 | 7.8% | [5.3%,11.4%] | 2.05 | (1.17 - 3.58) | - | - | 357, 372 | 9.00% | [7.1%,11.3%] | 2.46 | (1.54 - 3.93) | - | - | 778, 614 |  |
| 30-34 | 9.4% | [6.0%,14.3%] | 2.51 | (1.37 - 4.60) | - | - | 243, 287 | 6.90% | [5.0%,9.5%] | 1.85 | (1.10 - 3.11) | - | - | 596, 617 |  |
| 35-44 | 4.0% | [2.8%,5.6%] | 1.00 |  | - | - | 709, 815 | 3.90% | [2.6%,5.7%] | 1.00 |  | - | - | 742, 1315 |  |
| **Region** |  |  | **p=0.14** |  | **p=0.14** |  |  |  |  | **p=0.13** |  | **p=0.13** |  |  | **p=0.67** |
| England/Wales | 6.8% | [5.6%,8.3%] | 1.00 |  | 1.00 |  | 1529, 1703 | 6.20% | [5.3%,7.2%] | 1.00 |  | 1.00 |  | 2933, 3082 |  |
| Scotland | 3.4% | [1.4%,8.4%] | 0.49 | (0.19 - 1.28) | 0.48 | (0.18 - 1.27) | 139, 165 | 4.00% | [2.3%,6.9%] | 0.63 | (0.34 - 1.15) | 0.63 | (0.34 - 1.14) | 265, 285 |  |
| **Rurality** |  |  | **p=0.03** |  | **p=0.03** |  |  | - | - | - | - | - | - | - | - |
| Urban | 7.1% | [5.7%,8.8%] | 1.00 |  | 1.00 |  | 1243, 1393 | - | - | - | - | - | - | - | - |
| Rural | 2.2% | [0.8%,6.0%] | 0.29 | (0.10 - 0.86) | 0.3 | (0.10 - 0.87) | 132, 155 | - | - | - | - | - | - | - | - |
| **Ethnicity** |  |  | **p<0.001** |  | **p<0.001** |  |  |  |  | **p<0.001** |  | **p=0.0002** |  |  | **p=0.64** |
| White ^1^ | 5.9% | [4.7%,7.3%] | 1.00 |  | 1.00 |  | 1359, 1507 | 5.50% | [4.7%,6.5%] | 1.00 |  | 1.00 |  | 2785, 2844 |  |
| Mixed, multiple, or other ^2^ | 23.2% | [12.0%,40.3%] | 4.86 | (2.11 - 11.20) | 4.39 | (1.93 - 9.94) | 59, 71 | 14.00% | [7.7%,24.0%] | 2.78 | (1.40 - 5.54) | 2.65 | (1.31 - 5.33) | 109, 111 |  |
| Asian or Asian British ^3^ | 2.7% | [1.3%,5.6%] | 0.45 | (0.20 - 0.99) | 0.41 | (0.19 - 0.91) | 149, 184 | 3.30% | [1.6%,6.8%] | 0.58 | (0.27 - 1.27) | 0.60 | (0.28 - 1.32) | 190, 279 |  |
| Black or Black British ^4^ | 15.3% | [8.2%,26.7%] | 2.90 | (1.38 - 6.08) | 2.50 | (1.17 - 5.34) | 65, 72 | 15.60% | [9.5%,24.5%] | 3.17 | (1.76 - 5.73) | 3.23 | (1.78 - 5.86) | 108, 126 |  |
| **Sexual identity** |  |  | **p<0.001** |  | **p<0.001** |  |  |  |  | **p<0.001** |  | **p<0.001** |  |  | **p=0.37** |
| Heterosexual/straight | 5.9% | [4.7%,7.4%] | 1.00 |  | 1.00 |  | 1432, 1766 | 5.30% | [4.5%,6.3%] | 1.00 |  | 1.00 |  | 3074, 3255 |  |
| Gay or lesbian | 24.5% | [17.3%,33.4%] | 5.16 | (3.14 - 8.47) | 5.13 | (3.09 - 8.53) | 121, 49 | 36.00% | [24.3%,49.5%] | 9.97 | (5.54 - 17.91) | 9.68 | (5.36 - 17.49) | 82, 69 |  |
| Bisexual | 9.5% | [4.3%,19.3%] | 1.66 | (0.70 - 3.95) | 1.32 | (0.54 - 3.20) | 74, 20 | 7.10% | [2.1%,21.5%] | 1.35 | (0.37 - 4.91) | 1.29 | (0.36 - 4.60) | 31, 33 ^***^ |  |
| Other | - | - | - | - | - | - | 22, 12 ^**^ | - | - | - | - | - | - | 8, 6 ^**^ |  |
| **Social grade** |  |  | **p=0.01** |  | **p=0.01** |  |  | - | - | - | - | - | - | - | - |
| AB Higher and intermediate managerial/administrative/professional occupation | 9.5% | [7.2%,12.5%] | 1.00 |  | 1.00 |  | 583, 435 | - | - | - | - | - | - | - | - |
| C1 Supervisory, clerical and junior managerial/administrative/professional occupations/C2 Skilled manual occupations | 4.9% | [3.5%,6.9%] | 0.50 | (0.31 - 0.79) | 0.50 | (0.31 - 0.80) | 691, 1006 | - | - | - | - | - | - | - | - |
| D Semi-skilled and unskilled manual occupations/E On state benefit, unemployed and lowest grade occupations | 7.3% | [4.9%,10.7%] | 0.75 | (0.44 - 1.26) | 0.70 | (0.41 - 1.17) | 394, 426 | - | - | - | - | - | - | - | - |
| **Highest education qualification** |  |  | **p=0.01** |  | **p=0.03** |  |  |  |  | **p=0.08** |  | **p=0.04** |  |  | **p=0.07** |
| Degree | 6.40% | [4.8%,8.4%] | 1.00 |  | 1.00 |  | 813, 846 | 7.50% | [5.8%,9.6%] | 1.00 |  | 1.00 |  | 868, 1018 |  |
| Below degree | 5.60% | [4.1%,7.6%] | 0.87 | (0.56 - 1.35) | 0.80 | (0.51 - 1.27) | 750, 901 | 5.50% | [4.5%,6.6%] | 0.71 | (0.51 - 0.99) | 0.66 | (0.48 - 0.92) | 2162, 2152 |  |
| No qualification | 14.60% | [8.5%,24.1%] | 2.52 | (1.27 - 4.99) | 2.12 | (1.03 - 4.37) | 106, 121 | 3.80% | [1.4%,9.5%] | 0.48 | (0.17 - 1.35) | 0.50 | (0.18 - 1.41) | 163, 191 |  |
| **Born outside the UK** |  |  | **p=0.28** |  | **p=0.33** |  |  | - | - | - | - | - | - | - | - |
| No | 6.8% | [5.5%,8.3%] | 1.00 |  | 1.00 |  | 1492, 1678 | - | - | - | - | - | - | - | - |
| Yes | 4.6% | [2.2%,9.0%] | 0.65 | (0.31 - 1.40) | 0.68 | (0.32 - 1.47) | 160, 174 | - | - | - | - | - | - | - | - |
| **Relationship status** |  |  | **p=0.82** |  | **p=0.98** |  |  |  |  | **p<0.001** |  | **p=0.003** |  |  | **p=0.16** |
| Married/steady and living together | 6.30% | [4.8%,8.2%] | 1.00 |  | 1.00 |  | 975, 1098 | 4.30% | [3.3%,5.5%] | 1.00 |  | 1.00 |  | 1535, 2045 |  |
| Steady not living together | 7.60% | [4.2%,13.2%] | 1.22 | (0.62 - 2.41) | 0.97 | (0.46 - 2.05) | 153, 174 | 9.00% | [6.8%,11.8%] | 2.22 | (1.52 - 3.24) | 2.06 | (1.29 - 3.29) | 640, 504 |  |
| Not in a steady relationship | 6.90% | [4.9%,9.5%] | 1.10 | (0.70 - 1.73) | 0.95 | (0.59 - 1.54) | 524, 574 | 8.10% | [6.4%,10.2%] | 1.98 | (1.38 - 2.84) | 1.86 | (1.24 - 2.79) | 1006, 802 |  |
| **Days drinking, past 7 days** |  |  | **p=0.002** |  | **p=0.001** |  |  | - | - | - | - | - | - | - | - |
| 0 days | 5.4% | [3.6%,7.9%] | 1.00 |  | 1.00 |  | 509, 563 | - | - | - | - | - | - | - | - |
| 1-2 days | 5.1% | [3.6%,7.1%] | 0.94 | (0.54 - 1.63) | 0.91 | (0.52 - 1.58) | 667, 777 | - | - | - | - | - | - | - | - |
| 3-4 days | 12.0% | [8.6%,16.6%] | 2.42 | (1.38 - 4.22) | 2.41 | (1.38 - 4.23) | 324, 348 | - | - | - | - | - | - | - | - |
| 5-7 days | 6.0% | [3.2%,10.8%] | 1.12 | (0.52 - 2.41) | 1.15 | (0.54 - 2.48) | 161, 171 | - | - | - | - | - | - | - | - |
| **Currently smoker** |  |  | **p<0.001** |  | **p<0.001** |  |  |  |  | **p=0.03** |  | **p=0.05** |  |  | **p<0.001** |
| No | 3.30% | [2.5%,4.4%] | 1.00 |  | 1.00 |  | 1132, 1253 | 5.30% | [4.4%,6.5%] | 1.00 |  | 1.00 |  | 2086, 2290 |  |
| Yes | 12.90% | [10.0%,16.5%] | 4.33 | (2.86 - 6.57) | 4.10 | (2.69 - 6.26) | 520, 600 | 7.40% | [5.9%,9.1%] | 1.41 | (1.04 - 1.91) | 1.36 | (1.00 - 1.83) | 1112, 1076 |  |
| **Importance of sexual health, past year** |  |  | **p=0.002** |  | **p=0.002** |  |  | - | - | - | - | - | - | - | - |
| Very important/somewhat important | 8.2% | [6.6%,10.2%] | 1.00 |  | 1.00 |  | 1035, 1178 | - | - | - | - | - | - | - | - |
| Not very important/not important | 3.4% | [2.1%,5.4%] | 0.39 | (0.23 - 0.67) | 0.38 | (0.22 - 0.67) | 495, 541 | - | - | - | - | - | - | - | - |
| This does not apply to me | 2.6% | [0.5%,11.2%] | 0.29 | (0.06 - 1.42) | 0.33 | (0.07 - 1.63) | 108, 113 | - | - | - | - | - | - | - | - |
| **Symptoms of depression (PHQ-2) ^5^** |  |  | **p=0.001** |  | **p=0.005** |  |  |  |  | **p=0.27** |  | **p=0.28** |  |  | **p=0.25** |
| No | 4.7% | [3.5%,6.3%] | 1.00 |  | 1.00 |  | 1004, 1128 | 5.80% | [4.9%,6.9%] | 1.00 |  | 1.00 |  | 2862, 3038 |  |
| Yes | 9.1% | [6.9%,11.8%] | 2.04 | (1.33 - 3.14) | 1.88 | (1.21 - 2.93) | 627, 702 | 7.50% | [5.0%,11.1%] | 1.31 | (0.82 - 2.08) | 1.29 | (0.81 - 2.05) | 330, 323 |  |
| **Symptoms of anxiety (GAD-2) ^5^** |  |  | **p=0.001** |  | **p=0.006** |  |  | - | - | - | - | - | - | - | - |
| No | 5.0% | [3.8%,6.6%] | 1.00 |  | 1.00 |  | 1065, 1192 | - | - | - | - | - | - | - | - |
| Yes | 9.5% | [7.2%,12.3%] | 1.98 | (1.30 - 3.02) | 1.82 | (1.19 - 2.79) | 573, 645 | - | - | - | - | - | - | - | - |
| **Total sexual partners, past year ^6^** |  |  | **p<0.001** |  | **p<0.001** |  |  |  |  | **p<0.001** |  | **p<0.001** |  |  | **p=0.02** |
| 0 partners | 4.1% | [2.6%,6.6%] | 1.00 |  | 1.00 |  | 413, 461 | 0.30% | [0.0%,2.4%] | 1.00 |  | 1.00 |  | 169, 158 |  |
| 1 partner | 4.3% | [3.1%,6.0%] | 1.05 | (0.57 - 1.92) | 1.12 | (0.60 - 2.10) | 921, 1054 | 4.20% | [3.4%,5.4%] | 13.44 | (1.81 - 99.93) | 13.52 | (1.82 - 100.59) | 2086, 1404 |  |
| 2+ partners | 19.3% | [14.3%,25.6%] | 5.54 | (3.00 - 10.22) | 5.37 | (2.90 - 9.92) | 221, 233 | 12.30% | [10.0%,15.0%] | 42.45 | (5.73 - 314.60) | 41.25 | (5.52 - 308.35) | 902, 762 |  |
| **New sexual partners, past year ^6^** |  |  | **p<0.001** |  | **p<0.001** |  |  |  |  | **p<0.001** |  | **p<0.001** |  |  | **p=0.43** |
| 0 partners | 3.5% | [2.5%,4.9%] | 1.00 |  | 1.00 |  | 1179, 1334 | 3.40% | [2.6%,4.4%] | 1.00 |  | 1.00 |  | 1945, 2325 |  |
| 1 partner | 10.8% | [7.4%,15.4%] | 3.31 | (1.93 - 5.67) | 3.16 | (1.82 - 5.50) | 226, 257 | 8.90% | [6.7%,11.8%] | 2.81 | (1.87 - 4.24) | 2.90 | (1.84 - 4.57) | 660, 547 |  |
| 2+ partners | 23.1% | [16.3%,31.7%] | 8.21 | (4.70 - 14.34) | 7.60 | (4.21 - 13.72) | 144, 149 | 15.50% | [12.3%,19.3%] | 5.28 | (3.61 - 7.73) | 5.45 | (3.58 - 8.30) | 548, 447 |  |
| **Condom-less sex with a new partner on first occasion, past year ^6^** |  |  | **p<0.001** |  | **p<0.001** |  |  | - | - | - | - | - | - | - | - |
| None | 4.4% | [3.3%,5.8%] | 1.00 |  | 1.00 |  | 1286, 1440 | - | - | - | - | - | - | - | - |
| At least one | 16.7% | [12.5%,21.9%] | 4.41 | (2.82 - 6.91) | 4.09 | (2.57 - 6.51) | 260, 298 | - | - | - | - | - | - | - | - |
| **Previous same-sex experience, past 5 years ^7^** |  |  | **p<0.001** |  | **p<0.001** |  |  |  |  | **p<0.001** |  | **p<0.001** |  |  | **p=0.69** |
| No | 4.9% | [3.8%,6.2%] | 1.00 |  | 1.00 |  | 1453, 1721 | 5.30% | [4.4%,6.2%] | 1.00 |  | 1.00 |  | 3071, 3256 |  |
| Yes | 29.1% | [21.1%,38.7%] | 8.02 | (4.88 - 13.19) | 7.68 | (4.68 - 12.59) | 183, 114 | 27.90% | [19.7%,38.0%] | 7.00 | (4.26 - 11.50) | 6.72 | (4.06 - 11.10) | 126, 109 |  |
| **Pregnant in past year or currently pregnant** |  |  |  |  |  |  |  |  |  |  |  |  |  |  | - |
| No | - | - | - | - | - | - | - | - | - | - | - | - | - | - | - |
| Yes | - | - | - | - | - | - | - | - | - | - | - | - | - | - | - |
| **Used an STI-related service, past year** |  |  | **p<0.001** |  | **p<0.001** |  |  |  |  | **p<0.001** |  | **p<0.001** |  |  | **p=0.05** |
| No | 3.50% | [2.6%,4.7%] | 1.00 |  | 1.00 |  | 1458, 1646 | 2.90% | [2.2%,3.7%] | 1.00 |  | 1.00 |  | 2884, 3112 |  |
| Yes | 39.70% | [31.4%,48.5%] | 17.93 | (11.19 - 28.71) | 17.24 | (10.46 - 28.42) | 160, 158 | 47.70% | [41.4%,54.1%] | 30.96 | (21.34 - 44.93) | 42.92 | (27.97 - 65.88) | 298, 234 |  |
| **Unmet need for condoms, past year** |  |  | **p<0.001** |  | **p<0.001** |  |  | - | - | - | - | - | - | - | - |
| No | 3.80% | [2.8%,5.1%] | 1.00 |  | 1.00 |  | 1262, 1403 | - | - | - | - | - | - | - | - |
| Yes | 22.40% | [17.1%,28.6%] | 7.27 | (4.63 - 11.42) | 6.82 | (4.18 - 11.14) | 237, 267 | - | - | - | - | - | - | - | - |

CI=confidence intervals. OR=odds ratio. aOR=age-adjusted odds ratio. PHQ-2=Patient Health Questionnaire (2 item). GAD-2=Generalized anxiety disorder (2 item)

^*^ Age adjusted

^†^ Men or women aged 18-44 who were sexually-experienced. Trans men and trans women are included in data for men and women, respectively. 15 women and 22 men in Natsal-COVID responded 'prefer not to say' to questions about chlamydia testing. 423 women and 229 men in Natsal-3 did not answer the question. These individuals are excluded from the denominator.

^**^ Unweighted denominator <30. Results not shown due to small denominator

^***^Unweighted denominator <50. Results should be interpreted with caution due to small denominator.

^1^ White includes all those who identify as White English, Welsh, Scottish, Northern Irish, British, Irish, Gypsy or Irish Traveller, or from any other White background.

^2^ Mixed ethnicity includes those who identify as White and Black African, White and Black Caribbean, White and Asian or any other mixed or multiple ethnic background.

^3^ Asian includes those who identify as Indian, Pakistani, Bangladeshi, Chinese or from any other Asian background

^4^ Black includes those who identify as African, Caribbean, or from any other Black background.

^5^ Participants were classified as having symptoms of depression or anxiety if they scored three or more on the patient health questionnaire two item (PHQ-2) or generalised anxiety disorder two item (GAD-2) scales

^6^ Includes both opposite-sex and same-sex partners

^7^ Same-sex experience defined as oral/anal/vaginal sex

**Supplementary table 3.** Variations in reporting a cervical cancer screening among eligible participants aged 25-59 years in the first year following the start of a national lockdown in Britain (23/03/2020) compared with Natsal-3 (2010-12, past three years)

|  | **All eligible participants (Described female at birth, aged 25-59 yrs)** | | | | | | | | | | | | | | |
| --- | --- | --- | --- | --- | --- | --- | --- | --- | --- | --- | --- | --- | --- | --- | --- |
|  | **Natsal COVID (fieldwork 2021)** | | | | | | | **Natsal-3 (fieldwork 2010-12)** | | | | | | |  |
|  | **Weighted %** | **95% CI** | **OR** | **95% CI** | **aOR ^*^** | **95% CI** | **Denominator** ^†^ **(unweighted, weighted)** | **Weighted %** | **95% CI** | **OR** | **95% CI** | **aOR ^*^** | **95% CI** | **Denominator** ^†^ **(unweighted, weighted)** | **Interaction terms between surveys p-value** |
| **All ages (25-59 years)** | 10.3% | [9.2%,11.5%] | - | - | - | - | 2949, 2837 | 70.6% | [70.6%,70.6%] | - | - | - | - | 5176, 4770 |  |
| **Age (years)** |  |  | **p<0.001** |  |  |  |  |  |  | **p<0.001** |  |  |  |  | **p=0.01** |
| 25-29 | 16.3% | [13.4%,19.8%] | 2.72 | (1.94 - 3.83) | - | - | 582, 473 | 71.7% | [71.7%,71.7%] | 1.29 | (1.09 -1.53) | - | - | 1381, 683 |  |
| 30-34 | 12.4% | [9.7%,15.9%] | 1.98 | (1.36 - 2.88) | - | - | 463, 385 | 75.6% | [75.6%,75.6%] | 1.58 | (1.30 - 1.91) | - | - | 1036, 656 |  |
| 35-44 | 11.2% | [9.1%,13.8%] | 1.77 | (1.26 - 2.48) | - | - | 816, 762 | 74.0% | [74.0%,74.0%] | 1.45 | (1.21 - 1.72) | - | - | 1187, 1424 |  |
| 45-59 | 6.7% | [5.3%,8.4%] | 1.00 |  | - | - | 1088, 1217 | 66.3% | [66.3%,66.3%] | 1.00 |  | - | - | 1572, 2007 |  |
| **Region** |  |  | **p=0.17** |  | **p=0.10** |  |  |  |  | **p=0.62** |  | **p=0.52** |  |  | **p=0.07** |
| England/Wales | 10.6% | [9.4%,11.8%] | 1.00 |  | 1.00 |  | 2720, 2599 | 70.5% | [70.5%,70.5%] | 1.00 |  | 1.00 |  | 4740, 4333 |  |
| Scotland | 7.5% | [4.6%,12.0%] | 0.68 | (0.40 - 1.17) | 0.64 | (0.38 - 1.09) | 229, 238 | 71.8% | [71.8%,71.8%] | 1.07 | (0.83 - 1.38) | 1.09 | (0.84 - 1.40) | 436, 437 |  |
| **Rurality** |  |  | **p=0.02** |  | **p=0.03** |  |  | - | - | - | - | - | - | - | - |
| Urban | 10.7% | [9.4%,12.2%] | 1.00 |  | 1.00 |  | 2117, 2047 | - | - | - | - | - | - | - | - |
| Rural | 6.9% | [4.9%,9.8%] | 0.62 | (0.42 - 0.93) | 0.65 | (0.43 - 0.97) | 435, 408 | - | - | - | - | - | - | - | - |
| **Ethnicity** |  |  | **p=0.003** |  | **p=0.01** |  |  |  |  | **p=0.30** |  | **p=0.12** |  |  | **p=0.10** |
| White ^1^ | 9.9% | [8.8%,11.2%] | 1.00 |  | 1.00 |  | 2644, 2475 | 71.1% | [71.1%,71.1%] | 1.00 |  | 1.00 |  | 4527, 4160 |  |
| Mixed, multiple, or other ^2^ | 29.0% | [17.2%,44.5%] | 3.70 | (1.86 - 7.34) | 3.26 | (1.58 - 6.73) | 79, 66 | 72.6% | [72.6%,72.6%] | 1.08 | (0.69 - 1.69) | 1.03 | (0.65 - 1.61) | 141, 112 |  |
| Asian or Asian British ^3^ | 10.4% | [6.3%,16.5%] | 1.05 | (0.60 - 1.82) | 0.92 | (0.52 - 1.63) | 151, 188 | 66.5% | [66.5%,66.5%] | 0.81 | (0.62 - 1.05) | 0.74 | (0.56 - 0.96) | 304, 297 |  |
| Black or Black British ^4^ | 9.0% | [3.5%,21.1%] | 0.90 | (0.33 - 2.45) | 0.75 | (0.28 - 2.05) | 52, 73 | 66.9% | [66.9%,66.9%] | 0.82 | (0.56 - 1.20) | 0.81 | (0.56 - 1.18) | 194, 191 |  |
| **Sexual identity** |  |  | **p=0.001** |  | **p=0.003** |  |  |  |  | **p=0.93** |  | **p=0.86** |  |  | **p=0.01** |
| Heterosexual/straight | 10.2% | [9.1%,11.5%] | 1.00 |  | 1.00 |  | 2663, 2706 | 70.7% | [70.7%,70.7%] | 1.00 |  | 1.00 |  | 4994, 4626 |  |
| Gay or lesbian | 3.3% | [1.2%,8.6%] | 0.30 | (0.11 - 0.83) | 0.23 | (0.08 - 0.67) | 68, 31 | 66.3% | [66.3%,66.3%] | 0.82 | (0.44 - 1.51) | 0.79 | (0.43 - 1.46) | 71, 60 |  |
| Bisexual | 21.7% | [13.9%,32.3%] | 2.43 | (1.39 - 4.24) | 1.63 | (0.97 - 2.74) | 147, 40 | 70.7% | [70.7%,70.7%] | 1.00 | (0.58 - 1.73) | 0.89 | (0.51 - 1.56) | 87, 61 |  |
| Other | 5.3% | [1.1%,21.7%] | 0.49 | (0.10 - 2.44) | 0.24 | (0.05 - 1.20) | 43, 27 ^***^ | - | - | - | - | - | - | 14, 13 ^**^ |  |
| **Social grade** |  |  | **p=0.87** |  | **p=0.83** |  |  | - | - | - | - | - | - | - | - |
| AB Higher and intermediate managerial/administrative/professional occupation | 9.8% | [7.8%,12.2%] | 1.00 |  | 1.00 |  | 742, 660 | - | - | - | - | - | - | - | - |
| C1 Supervisory, clerical and junior managerial/administrative/professional occupations/C2 Skilled manual occupations | 10.5% | [9.0%,12.2%] | 1.09 | (0.80 - 1.47) | 1.10 | (0.81 - 1.50) | 1553, 1554 | - | - | - | - | - | - | - | - |
| D Semi-skilled and unskilled manual occupations/E On state benefit, unemployed and lowest grade occupations | 10.3% | [8.2%,13.0%] | 1.07 | (0.74 - 1.53) | 1.06 | (0.73 - 1.53) | 654, 623 | - | - | - | - | - | - | - | - |
| **Highest education qualification** |  |  | **p=0.03** |  | **p=0.16** |  |  |  |  | **p<0.001** |  | **p<0.001** |  |  | **p=0.92** |
| Degree | 11.8% | [10.2%,13.7%] | 1.00 |  | 1.00 |  | 1488, 1393 | 75.00% | [75.0%,75.0%] | 1.00 |  | 1.00 |  | 1629, 1486 |  |
| Below degree | 8.9% | [7.5%,10.6%] | 0.73 | (0.56 - 0.94) | 0.81 | (0.63 - 1.05) | 1346, 1331 | 69.90% | [69.9%,69.9%] | 0.77 | (0.66 - 0.90) | 0.81 | (0.70 - 0.95) | 3070, 2838 |  |
| No qual | 7.5% | [3.9%,14.0%] | 0.60 | (0.29 - 1.23) | 0.62 | (0.30 - 1.27) | 115, 114 | 60.60% | [60.6%,60.6%] | 0.51 | (0.40 - 0.66) | 0.58 | (0.45 - 0.75) | 469, 438 |  |
| **Born outside the UK** |  |  | **p<0.001** |  | **p=0.001** |  |  | - | - | - | - | - | - | - | - |
| No | 9.3% | [8.2%,10.5%] | 1.00 |  | 1.00 |  | 2579, 2456 | - | - | - | - | - | - | - | - |
| Yes | 17.5% | [13.5%,22.3%] | 2.07 | (1.48 - 2.90) | 1.78 | (1.26 - 2.51) | 349, 358 | - | - | - | - | - | - | - | - |
| **Relationship status** |  |  | **p=0.05** |  | **p=0.15** |  |  |  |  | **p<0.001** |  | **p<0.001** |  |  | **p=0.27** |
| Married/steady and living together | 9.9% | [8.7%,11.4%] | 1.00 |  | 1.00 |  | 1917, 1871 | 71.7% | [71.7%,71.7%] | 1.00 |  | 1.00 |  | 3204, 3465 |  |
| Steady not living together | 15.4% | [11.0%,21.0%] | 1.64 | (1.09 - 2.48) | 1.47 | (0.97 - 2.25) | 227, 202 | 75.8% | [75.8%,75.8%] | 1.24 | (0.97 - 1.58) | 1.18 | (0.92 - 1.50) | 653, 409 |  |
| Not in a steady relationship | 10.0% | [7.9%,12.5%] | 1.01 | (0.75 - 1.35) | 0.95 | (0.70 - 1.28) | 797, 754 | 64.7% | [64.7%,64.7%] | 0.72 | (0.62 - 0.84) | 0.72 | (0.62 - 0.85) | 1287, 873 |  |
| **Days drinking, past 7 days** |  |  | **p=0.11** |  | **p=0.09** |  |  | - | - | - | - | - | - | - | - |
| 0 days | 9.3% | [7.8%,11.0%] | 1.00 |  | 1.00 |  | 1342, 1302 | - | - | - | - | - | - | - | - |
| 1-2 days | 11.9% | [10.0%,14.2%] | 1.33 | (1.00 - 1.75) | 1.31 | (0.99 - 1.73) | 1019, 972 | - | - | - | - | - | - | - | - |
| 3-4 days | 8.6% | [6.1%,12.0%] | 0.92 | (0.60 - 1.41) | 0.92 | (0.60 - 1.42) | 358, 346 | - | - | - | - | - | - | - | - |
| 5-7 days | 12.4% | [8.6%,17.7%] | 1.39 | (0.88 - 2.19) | 1.53 | (0.96 - 2.42) | 222, 208 | - | - | - | - | - | - | - | - |
| **Currently smoker** |  |  | **p=0.003** |  | **p=0.02** |  |  |  |  | **p<0.001** |  | **p<0.001** |  |  | **p=0.0003** |
| No | 9.4% | [8.3%,10.7%] | 1.00 |  | 1.00 |  | 2383, 2296 | 72.4% | [72.4%,72.4%] | 1.00 |  | 1.00 |  | 3756, 3622 |  |
| Yes | 14.0% | [11.2%,17.4%] | 1.57 | (1.17 - 2.10) | 1.42 | (1.05 - 1.92) | 557, 532 | 64.9% | [64.9%,64.9%] | 0.70 | (0.61 - 0.82) | 0.68 | (0.59 - 0.79) | 1420, 1148 |  |
| **Importance of sexual health, past year** |  |  | **p<0.001** |  | **p<0.001** |  |  | - | - | - | - | - | - | - | - |
| Very important/somewhat important | 13.3% | [11.6%,15.2%] | 1.00 |  | 1.00 |  | 1520, 1428 | - | - | - | - | - | - | - | - |
| Not very important/not important | 8.3% | [6.7%,10.2%] | 0.59 | (0.45 - 0.78) | 0.67 | (0.51 - 0.89) | 1050, 1032 | - | - | - | - | - | - | - | - |
| This does not apply to me | 4.0% | [2.2%,7.0%] | 0.27 | (0.14 - 0.50) | 0.30 | (0.16 - 0.56) | 303, 298 | - | - | - | - | - | - | - | - |
| **Symptoms of depression (PHQ-2) ^5^** |  |  | **p=0.21** |  | **p=0.93** |  |  |  |  | **p=0.02** |  | **p=0.03** |  |  | **p=0.99** |
| No | 9.8% | [8.6%,11.2%] | 1.00 |  | 1.00 |  | 2033, 1983 | 71.3% | [71.3%,71.3%] | 1.00 |  | 1.00 |  | 4535, 4210 |  |
| Yes | 11.4% | [9.4%,13.8%] | 1.18 | (0.91 - 1.55) | 1.01 | (0.77 - 1.34) | 885, 826 | 65.9% | [65.9%,65.9%] | 0.78 | (0.63 - 0.96) | 0.80 | (0.64 - 0.98) | 629, 546 |  |
| **Symptoms of anxiety (GAD-2) ^5^** |  |  | **p=0.07** |  | **p=0.54** |  |  | - | - | - | - | - | - | - | - |
| No | 9.6% | [8.4%,11.1%] | 1.00 |  | 1.00 |  | 1976, 1955 | - | - | - | - | - | - | - | - |
| Yes | 11.9% | [9.9%,14.2%] | 1.26 | (0.98 - 1.64) | 1.09 | (0.84 - 1.41) | 956, 866 | - | - | - | - | - | - | - | - |
| **Total sexual partners, past year ^6^** |  |  | **p<0.001** |  | **p=0.006** |  |  |  |  | **p<0.001** |  | **p<0.001** |  |  | **p=0.01** |
| 0 partners | 8.4% | [6.5%,10.6%] | 1.00 |  | 1.00 |  | 873, 860 | 58.0% | [58.0%,58.0%] | 1.00 |  | 1.00 |  | 634, 543 |  |
| 1 partner | 10.8% | [9.4%,12.4%] | 1.33 | (0.98 - 1.80) | 1.28 | (0.94 - 1.75) | 1792, 1704 | 72.3% | [72.3%,72.3%] | 1.89 | (1.56 - 2.29) | 1.72 | (1.41 - 2.09) | 3884, 3762 |  |
| 2+ partners | 24.4% | [16.8%,34.2%] | 3.55 | (2.06 - 6.10) | 2.48 | (1.42 - 4.33) | 114, 99 | 73.1% | [73.1%,73.1%] | 1.97 | (1.47 - 2.63) | 1.64 | (1.22 - 2.21) | 579, 385 |  |
| **New sexual partners, past year ^6^** |  |  | **p<0.001** |  | **p=0.002** |  |  |  |  | **p=0.12** |  | **p=0.22** |  |  | **p=0.06** |
| 0 partners | 9.9% | [8.8%,11.2%] | 1.00 |  | 1.00 |  | 2560, 2469 | 70.5% | [70.5%,70.5%] | 1.00 |  | 1.00 |  | 4211, 4089 |  |
| 1 partner | 13.9% | [9.1%,20.7%] | 1.47 | (0.89 - 2.42) | 1.20 | (0.73 - 1.97) | 163, 148 | 69.5% | [69.5%,69.5%] | 0.95 | (0.76 - 1.19) | 0.88 | (0.70 - 1.10) | 620, 435 |  |
| 2+ partners | 32.3% | [19.7%,48.1%] | 4.33 | (2.20 - 8.55) | 3.29 | (1.69 - 6.40) | 51, 43 | 77.5% | [77.5%,77.5%] | 1.44 | (1.00 - 2.06) | 1.24 | (0.87 - 1.78) | 261, 160 |  |
| **Condom-less sex with a new partner on first occasion, past year ^6^** |  |  | **p<0.001** |  | **p=0.01** |  |  | - | - | - | - | - | - | - | - |
| None | 10.0% | [8.9%,11.3%] | 1.00 |  | 1.00 |  | 2658, 2550 | - | - | - | - | - | - | - | - |
| At least one | 20.5% | [13.9%,29.1%] | 2.31 | (1.42 - 3.75) | 1.86 | (1.14 - 3.03) | 131, 120 | - | - | - | - | - | - | - | - |
| **Previous same-sex experience, past 5 years ^7^** |  |  | **p=0.41** |  | **p=0.26** |  |  |  |  | **p=0.56** |  | **p=0.86** |  |  | **p=0.21** |
| No | 10.4% | [9.3%,11.7%] | 1.00 |  | 1.00 |  | 2761, 2707 | 70.5% | [70.5%,70.5%] | 1.00 |  | 1.00 |  | 4965, 4606 |  |
| Yes | 8.1% | [4.4%,14.4%] | 0.76 | (0.39 - 1.47) | 0.68 | (0.35 - 1.32) | 127, 65 | 72.8% | [72.8%,72.8%] | 1.12 | (0.77 - 1.61) | 1.03 | (0.71 - 1.50) | 209, 161 |  |
| **Chlamydia test, past year** |  |  | **p<0.001** |  | **p<0.001** |  |  |  |  | **p<0.001** |  | **p<0.001** |  |  | **p<0.001** |
| No/not sure | 9.3% | [8.3%,10.5%] | 1.00 |  | 1.00 |  | 2820, 2724 | 72.7% | [72.7%,72.7%] | 1.00 |  | 1.00 |  | 2840, 2277 |  |
| Yes | 40.6% | [30.8%,51.2%] | 6.62 | (4.23 - 10.36) | 2.65 | (1.64 - 4.28) | 105, 89 | 81.5% | [81.5%,81.5%] | 1.65 | (1.29 - 2.12) | 1.68 | (1.30 - 2.18) | 702, 433 |  |
| **HIV test, past year** |  |  | **p<0.001** |  | **p<0.001** |  |  |  |  | **p=0.28** |  | **p=0.79** |  |  | **p<0.001** |
| No/not sure | 9.5% | [8.5%,10.7%] | 1.00 |  | 1.00 |  | 2783, 2680 | 70.4% | [70.4%,70.4%] | 1.00 |  | 1.00 |  | 4476, 4214 |  |
| Yes | 26.3% | [18.8%,35.5%] | 3.38 | (2.15 - 5.31) | 5.05 | (3.19 - 7.99) | 145, 135 | 73.7% | [73.7%,73.7%] | 1.18 | (0.87 - 1.60) | 0.96 | (0.71 - 1.31) | 361, 243 |  |
| **Used an STI-related service, past year** |  |  | **p<0.001** |  | **p<0.001** |  |  |  |  | **p=0.001** |  | **p=0.02** |  |  | **p=0.002** |
| No | 9.2% | [8.2%,10.4%] | 1.00 |  | 1.00 |  | 2830, 2730 | 70.6% | [70.6%,70.6%] | 1.00 |  | 1.00 |  | 4856, 4542 |  |
| Yes | 38.0% | [28.9%,48.1%] | 6.04 | (3.92 - 9.30) | 4.74 | (3.02 - 7.44) | 119, 107 | 82.9% | [82.9%,82.9%] | 2.01 | (1.32 - 3.06) | 1.65 | (1.08 - 2.53) | 227, 141 |  |
| **Unmet need for condoms, past year** |  |  | **p=0.20** |  | **p=0.95** |  |  | - | - | - | - | - | - | - | - |
| No | 10.70% | [9.5%,12.0%] | 1.00 |  | 1.00 |  | 2525, 2410 | - | - | - | - | - | - | - | - |
| Yes | 15.10% | [8.8%,24.7%] | 1.50 | (0.80 - 2.80) | 1.02 | (0.53 - 1.96) |  | - | - | - | - | - | - | - | - |

CI=confidence intervals. OR=odds ratio. aOR=age-adjusted odds ratio. PHQ-2=Patient Health Questionnaire (2 item). GAD-2=Generalized anxiety disorder (2 item)

^*^ Age adjusted

^†^ Participants described female at birth aged 25-29. 90 eligible participants responded 'prefer not to say' to questions about cervical cancer screening. 144 in Natsal-3 did not answer the question. These individuals are excluded from the denominator.

^**^ Unweighted denominator <30. Results not shown due to small denominator

^***^Unweighted denominator <50. Results should be interpreted with caution due to small denominator.

^1^ White includes all those who identify as White English, Welsh, Scottish, Northern Irish, British, Irish, Gypsy or Irish Traveller, or from any other White background.

^2^ Mixed ethnicity includes those who identify as White and Black African, White and Black Caribbean, White and Asian or any other mixed or multiple ethnic background.

^3^ Asian includes those who identify as Indian, Pakistani, Bangladeshi, Chinese or from any other Asian background

^4^ Black includes those who identify as African, Caribbean, or from any other Black background.

^5^ Participants were classified as having symptoms of depression or anxiety if they scored three or more on the patient health questionnaire two item (PHQ-2) or generalised anxiety disorder two item (GAD-2) scales

^6^ Includes both opposite-sex and same-sex partners

^7^ Same-sex experience defined as oral/anal/vaginal sex

All percentages are weighted. These are row percentages which describe reported use of cervical cancer screening in the past year (Natsal-COVID) or past three years (Natsal-3) within certain subgroups.
